# Whole-genome sequencing implicates rare, low-frequency and structural non-coding variation at the *SCN5A* locus in Brugada syndrome

**DOI:** 10.64898/2026.07.07.26356386

**Authors:** Alex Lipov, Manon Baudic, Pierre Lindenbaum, Isabella Mengarelli, Matthew J. O’Neill, Fernanda M. Bosada, Yanushi Wijeyeratne, Luis de la Higuera Romero, Maarten Kooyman, Marion Gaudin, Graziella Aquilina, Leander Beekman, Estelle Baron, Mathilde Bertrand, Zoya Kingsbury, Mark T. Ross, Marre Corver, Paola Lombardi, Ingrid Krapels, Paul G. Volders, Rafik Tadros, Fenna Tuijnenburg, Karel van Duijvenboden, Ammar Al-Chalabi, Jan H. Veldink, Sean J. Jurgens, Aurélie Thollet, Eric Charpentier, Camille Maiano, Philippe Mabo, Antoine Leenhardt, Frederic Sacher, Arjan C. Houweling, Hanno L. Tan, Vincent M. Christoffels, Michael W. Tanck, Andrew Grace, Koonlawee Nademanee, Apichai Khongphatthanayothin, Andrew M. Glazer, Jean François Deleuze, FranceGenRef consortium, Juan Pablo Ochoa, Jérôme Montnach, Michel De Waard, Pieter G. Postema, Ahmad S. Amin, Jean-Baptiste Gourraud, Pascale Guicheney, Dan M. Roden, Jean-Jacques Schott, Christian Dina, Vincent Probst, Pier D. Lambiase, Elijah R. Behr, Arthur A.M. Wilde, Richard Redon, Roddy Walsh, Julien Barc, Connie R. Bezzina

## Abstract

Brugada syndrome (BrS) is an inherited cardiac condition characterized by a hallmark ECG pattern and an increased risk of sudden cardiac death. Central to the aetiology of BrS, the *SCN5A* region harbours both common non-coding risk variants and rare coding variants that are causative in approximately 20% of patients. However, rare non-coding genetic variation in this region remains largely unexplored. Here, we used whole-genome sequencing (WGS) of 752 European-ancestry BrS cases and 1,827 ancestry-matched controls to identify BrS-associated rare non-coding genetic variation at the *SCN5A* locus. Sliding-window and *cis*-regulatory element (CRE)-based rare-variant aggregate testing implicated three conserved CREs, including a dense aggregation of case singleton variants within a 178 bp enhancer in intron 17 of *SCN5A* which replicated in an independent BrS cohort. Prioritised BrS-associated rare and low-frequency non-coding variants within these elements were predicted to alter cardiac transcription factor motifs, and altered CRE activity in hiPSC-CM luciferase assays or were associated with BrS-relevant ECG endophenotypes in the UK Biobank. Single-variant analysis across the region identified a Bonferroni-significant five-fold case-enriched low-frequency variant within a known CRE in intron 1 of *SCN5A,* which replicated, was associated with slower cardiac conduction in the UK Biobank and accounted for part of the BrS GWAS signal at this locus. Structural variant analyses identified a 10.5 kb deletion upstream of *SCN5A* in a BrS case that encompassed a cardiac CRE and reduced sodium current density in a hiPSC-CM model, as well as a 6 kb BrS-enriched retrotransposon insertion in *SCN5A* that appeared to underlie part of the GWAS signal in this region. Together, these findings implicate rare and low-frequency non-coding variation at the *SCN5A* locus in BrS susceptibility and demonstrate the value of targeted WGS analysis of key disease loci.

## Introduction

Brugada syndrome (BrS) is an inherited cardiac condition characterised by the presence of ST-segment elevation and T-wave inversion in the right precordial ECG leads with a concomitant increased risk of sudden cardiac death^1–3^. Approximately 20% of BrS cases of European ancestry have a rare loss-of-function coding variant in *SCN5A*, which encodes the alpha subunit of the cardiac voltage-gated sodium channel^4^. However, the genetic aetiology in the other 80% of cases is unknown. Although rare coding variants in at least 20 other genes have been implicated in BrS, *SCN5A* is the only gene classified by the ClinGen Gene Curation Expert Panel as having definitive evidence for a causal association with the condition^5^. Genome-wide association studies (GWAS) have also identified common non-coding variants which predispose individuals to BrS, with the largest-to-date European-ancestry study identifying 12 associated loci across the genome^6^. Out of the 21 conditionally-independent genome-wide significant SNPs across these loci, eight were located in the vicinity of *SCN5A,* highlighting the primacy of this genomic region in BrS (Figure 1A). However, rare non-coding genetic variation at this locus remains largely unexplored in BrS.

**Figure 1.**
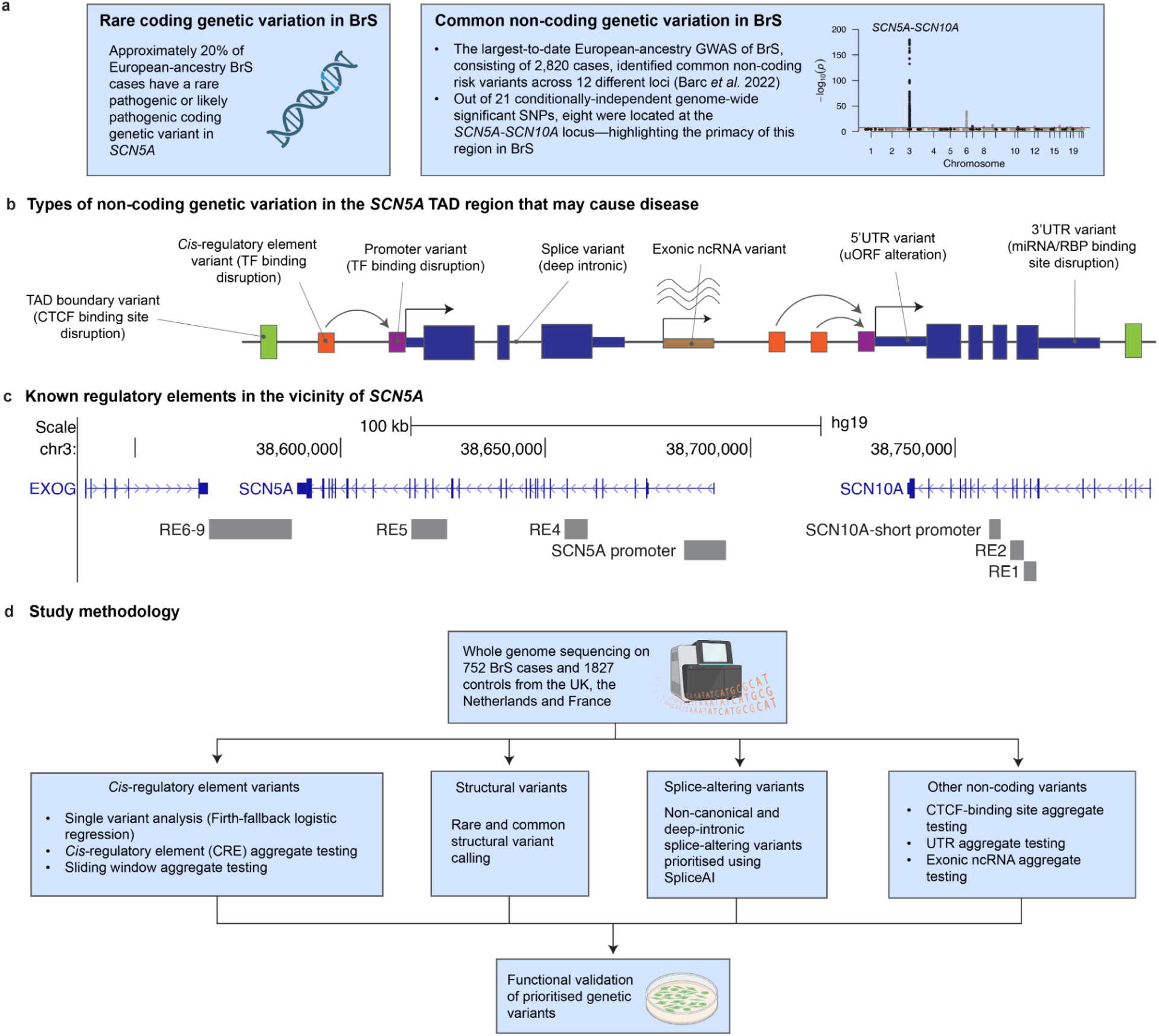
| Study background and methodology. (A) The current status of BrS genetics; rare non-coding variation has been largely unexplored. (B) The various mechanisms by which non-coding variants can cause disease. (C) The locations of previously characterised regulatory elements in the SCN5A–SCN10A region. (D) Overview of the study design for identifying and validating various classes of non-coding variants in the SCN5A region using WGS of 752 BrS cases and 1,827 controls. SNP, single-nucleotide polymorphism; BrS, Brugada syndrome; TAD, topologically-associating domain; RBP, RNA-binding protein; CRE, cis-regulatory element; TF, transcription factor; uORF, upstream open reading frame; CTCF, CCCTC-binding factor.

Given that ∼97% of genetic variation is rare^7^ and ∼98% of the human genome is non-coding^8^, the vast majority of genetic variation consists of rare non-coding variants. Moreover, under purifying selection, deleterious variants are maintained at low population frequencies, a principle well established for protein-coding variation, where lower minor allele frequency is associated with larger effect sizes. Emerging evidence suggests a similar relationship for non-coding variation, whereby, among variants with deleterious effects, rare variants tend to have higher effect sizes than their common counterparts and thus are potentially of greater clinical relevance^9,10^.

However, identifying rare deleterious non-coding variants remains challenging, as low minor allele counts limit statistical power and the large search space of the incompletely annotated non-coding genome complicates variant aggregation efforts. Simultaneously, experimental validation of these variants remains challenging and laborious. Rare non-coding variants can cause disease through a variety of mechanisms; the major ones are illustrated in Figure 1B. One important class of non-coding variation is that which disrupts transcription factor (TF) binding in a *cis*-regulatory element. The major regulatory elements of the *SCN5A-SCN10A* region have been characterized in prior work (RE1-9, Figure 1C)^11–13^. Several of the BrS GWAS signals overlap these regulatory elements, and we previously identified and functionally validated a recurrent loss-of-function variant within RE5 that is causal in 4% of BrS patients from Thailand,^14^ where the condition is particularly prevalent.

In this work, we performed whole-genome sequencing (WGS) on 752 BrS cases from the UK, the Netherlands and France, along with 1,827 ancestry-matched controls in order to identify rare and low-frequency non-coding genetic variation in the *SCN5A* region that explains a proportion of the missing genetic aetiology of BrS (Figure 1D).

## Methods

### Study population

WGS was performed on BrS cases and ancestry-matched controls from the UK, the Netherlands and France (*n* = 752 cases and *n* = 1,827 controls, after sample QC). Dutch cases (*n* = 262) were ascertained through Amsterdam University Medical Center (Amsterdam UMC) and Maastricht University Medical Center+ (Maastricht UMC+); UK cases (*n* = 147) through St George’s University Hospitals NHS Foundation Trust and University College London (UCL); and French cases (*n* = 343) through the national healthcare network for inherited cardiac diseases (Cardiogen), which includes the university hospitals of Nantes, Rennes, Bordeaux and Paris.

BrS was diagnosed according to the 2013 HRS/EHRA/APHRS consensus statement^15^, the 2015 ESC guidelines^16^ and the 2017 AHA/ACC/HRS guidelines^17^. All cases exhibited a type 1 Brugada ECG pattern, with ≥ 2 mm coved ST-segment elevation in ≥ 1 right precordial lead (V1-V2), either spontaneously or following intravenous provocation with a class I antiarrhythmic drug. ECGs were centrally reviewed by an electrophysiologist with expertise in BrS. The clinical characteristics of the BrS cases are provided in Table S1.

UK and Dutch controls were obtained from Project MinE^18^; French controls were obtained from the FranceGenRef consortium. All participants provided written informed consent. The study was approved by the relevant ethics committees in France (Nantes: DC-2011-1399), the Netherlands (Amsterdam UMC METC: W20_226 #20.260) and the UK (St George’s: REC Ref: 10/H0803/121, IRAS number 53663; UCL: 15/LO/0549).

### Whole-genome sequencing

Region-restricted BAMs spanning the *SCN5A* TAD and flanking sequence (chr3:38.0–39.5 Mb; GRCh37) were used for joint variant calling across all samples. Variant– and sample-level quality control were applied (Supplemental Methods), leaving 25,162 variants and 2,579 samples. Ancestry principal components (PCs) were computed for the downstream regression models. Full details of the WGS pipeline are provided in the Supplemental Methods.

### Non-coding variant annotation

Variants were annotated using ANNOVAR^19^ and VEP^20^. Computationally predicted functional effects were obtained using RegVar^21^ (3′UTR) and UTRannotator^22^ (5′UTR). Variants within candidate *cis*-regulatory elements (CREs) were evaluated for predicted TF motif disruption using motifbreakR^23^ and HOMER2^24^ (Supplemental Methods). Rare variants predicted to alter *SCN5A* splicing, excluding those at canonical splice sites, were prioritised using SpliceAI^25^ (criteria detailed in Supplemental Methods).

### Defining non-coding regions of interest

Cardiac epigenomic datasets were used to define regions of interest within the *SCN5A* TAD (chr3:38488336-39139033; GRCh37). Twenty-five conserved, cardiac-accessible chromatin elements (E1–E25), hereafter referred to as candidate CREs, and 18 CTCF-binding regions (C1–C18) were defined based on evolutionary conservation and cardiac epigenomic datasets (Supplemental Methods; Tables S2–S3). More concretely, candidate CREs were defined as regions that were evolutionarily conserved (100 Vert. Cons. PhyloP), and had either a human left atrial (LA) ATAC-seq^26^ or a human ventricular-like cardiomyocyte (VLCM) ATAC-seq^27^ peak. For each of these candidate CRE regions, ChIP-seq signals were considered; elements with only a CTCF ChIP-seq peak were excluded. Full details can be found in the Supplemental Methods.

### Association analyses

Genetic association analyses were performed using REGENIE^28^. In step-one, a genome-wide model was fitted using common variants; this was used to account for relatedness and population structure in downstream step-two analyses (Supplemental Methods). The covariates in step-one were sex, national centre, *SCN5A* rare-variant carrier status and the first 10 ancestry PCs. Where appropriate, we conditioned on the 2022 BrS GWAS SNPs in step-two analyses to account for the common variant signal (Supplemental Methods). Step-two analyses were performed as detailed below.

Burden-based aggregate testing of singleton variants (minor allele count, MAC = 1) was performed for the 25 candidate CREs. Regions with a cumulative minor allele count (cMAC) ≥ 5 were retained for analysis, resulting in 16 tested CREs. A Bonferroni correction was applied to account for multiple testing (*α*_b_ = 0.05/16). A similar approach was employed for the 18 CTCF-binding regions (*α*_b_ = 0.05/13, for the 13 regions that passed cMAC ≥ 5).

Sliding-window burden testing (200 bp windows, each overlapping adjacent windows by 20%) was conducted across the *SCN5A* TAD, including only singleton variants with CADD > 10 and requiring cMAC ≥ 5. Of 4,066 windows, 15 met the criteria for testing (*α*_b_ = 0.05/15).

Single-variant testing of variants with AAF_gnomAD-popmax_ < 1% across the *SCN5A* region was performed using Firth-fallback logistic regression (*α*_b_ = 0.05/18,432). Conditional analyses were performed on select variants by repeating this step-two model with the variant of interest included as a covariate.

### Structural variants

Structural variants (SVs) were identified using Delly2^29^, Manta^30^ and IndexCov^31^. Targeted SV calling was performed on a common *SCN5A* mobile element insertion (MEI) using MELT^32^. Conditional analyses were performed between the MEI and the previously reported conditionally independent genome-wide significant BrS GWAS SNPs by repeating a single-variant analysis with and without conditioning on the MEI.

### Genetic replication in an independent BrS cohort

We sought to replicate the identified genetic associations in an independent BrS referral cohort that underwent genetic testing by next-generation sequencing at Health in Code S.L., Spain. Individuals referred for BrS genetic testing (483 probands) and individuals with non-channelopathy cardiovascular conditions (12,259 probands) were selected as cases and non-BrS controls respectively. Notably, the panel assay used for these samples incorporates all *SCN5A* introns, enabling the interrogation of non-coding variants in these regions (though not for *SCN10A* introns). See Supplemental Methods for further details.

### Association analysis with ECG parameters in the UK Biobank

To support the role of prioritised rare non-coding variants on BrS risk, we sought evidence for effects on electrocardiogram (ECG) conduction parameters of relevance to BrS in the UK Biobank (UKB). To this end, we performed a genome-wide association analysis for PR (*n* = 110,723 individuals) and QRS intervals (*n* = 112,020 individuals) in European ancestry individuals. We tested all SNVs and small indels with MAC > 10 within the prioritised candidate CREs in the *SCN5A*–*SCN10A* region (E14, E17, E22 and E23) for association with these endophenotypes. The Supplemental Methods provide further methodological detail.

### Functional validation

Prioritised variants were functionally evaluated using three distinct experimental approaches. First, variants within candidate CREs were assessed using luciferase reporter assays in human induced pluripotent stem cell-derived cardiomyocytes (hiPSC-CMs). Second, a 10 kb deletion upstream of *SCN5A* identified in a BrS case was introduced into hiPSC-CMs using CRISPR-Cas9, and mutant and control isogenic cell lines were electrophysiologically characterised using automated high-throughput patch clamp. Third, selected splice variants were assessed using minigene assays and CRISPR-Cas9 editing of hiPSC-CMs. Detailed experimental procedures are provided in the Supplemental Methods.

## Results

### Cohort characteristics and protein-coding variants

The BrS cases had a mean age at diagnosis of 47.9 ± 13.2 years, 75% were male, 8.4% experienced ventricular tachycardia or ventricular fibrillation, and 33.4% exhibited a spontaneous type 1 BrS ECG pattern (Table S1). This BrS cohort largely overlaps the BrS cases included in the 2022 BrS GWAS^6^ and our earlier variant interpretation study^4^. As expected for a standard well-defined European-ancestry BrS cohort, 18.6% (*n* = 140) of cases carried a rare coding variant in *SCN5A*—defined as a nonsynonymous exonic or essential splice site variant with a gnomADv4 FAF_95-popmax_ < 1×10^-4^ (gnomAD exomes dataset; transcript ENST00000333535). Among controls, 2.3% were carriers (*n* = 42). Of the other protein-coding genes in the *SCN5A* TAD, the *SCN10A* gene has been previously implicated in BrS^6^. We found no statistically significant case enrichment of nonsynonymous coding variants (AAF_gnomAD-NFE_ < 1×10^-4^) in *SCN10A* (burden *p* = 0.74; 17/752 = 2.3% carriers in cases, 32/1827 = 1.8% carriers in controls) nor in the recently described *SCN10A*-short isoform comprising the last 7 exons of *SCN10A* and thought to modulate sodium channel function^12^ (burden *p* = 0.90; 2/752 = 0.3% carriers in cases, 8/1827 = 0.4% carriers in controls). Of the TAD genes, only *SCN5A* reached Bonferroni-corrected significance (burden *p* = 1.5e-40, *α*_b_ = 0.05/24).

### The contribution of non-canonical and deep-intronic splice-site variants

While *SCN5A* canonical splice-site variants have been well characterised, non-canonical and deep-intronic splice variants have been understudied. We therefore investigated their contribution to BrS.

### Non-canonical splice-altering variants near *SCN5A* intron-exon boundaries

Four variants located near *SCN5A* exon–intron junctions, but outside the canonical donor (+1/+2) and acceptor (−1/−2) splice-site positions, were each identified in a single case and predicted by SpliceAI^25^ to alter splicing. We previously functionally validated two of these variants: the synonymous variant c.1890G>A (p.Thr630Thr), which caused intron 10 retention in a minigene assay, and c.4437+5G>A, which caused a 32 bp exon extension and frameshift in CRISPR-Cas9-edited hiPSC-CMs^33^. Of the remaining two variants, the synonymous variant c.3963G>A (p.Arg1321Arg) was assessed here using a minigene assay and found to induce both exon skipping and pseudo-exon inclusion (Figure S6). The intronic variant c.3666+22G>T was predicted to cause exon extension and a frameshift, which were confirmed by analysis of RNA from the patient’s blood.

### Limited role of deep-intronic *SCN5A* splice variants

Deep-intronic splice-altering variants, which can cause exon extension or cryptic pseudo-exon inclusion, may explain a proportion of the unresolved genetic aetiology of BrS^34^ (for instance, deleterious *MYBPC3* deep intronic splice-altering variants have been reported in at least 2% of hypertrophic cardiomyopathy cases^35^). We used SpliceAI to identify variants that were predicted to create a cryptic exon (gain of both acceptor and donor sites) and were enriched, or only occurring, in cases. A single candidate variant was identified, c.2263-310G>T (AAF_cases_ = 0.60%, AAF_controls_ = 0.22%), but no effect on splicing was observed in CRISPR-Cas9 edited hiPSC-CMs (Figure S6). These findings suggest that deep intronic splice-altering variants in *SCN5A* do not appear to be a prevalent pathogenic variant class for BrS.

### Rare non-coding burden testing implicates candidate regulatory elements in the *SCN5A* TAD

We sought to identify BrS-associated rare non-coding variants within candidate CREs of the *SCN5A* region. Specifically, we considered the region chr3:38,488,336–39,139,033 (GRCh37), corresponding to the cardiomyocyte TAD harbouring *SCN5A*. This interval was defined based on cardiac-specific Hi-C data^36^, promoter capture Hi-C data^37^, and the positions of convergent CTCF-occupied sites^38^. Using genetic conservation and cardiac ATAC-seq data^26,27^, 25 conserved cardiac-specific accessible chromatin elements (E1-E25) were defined across the region (blue marks in Figure 2A; mean length 565 bp). These were considered candidate CREs henceforth. E1–E25 span a broader genomic region and provide a more finely delineated representation of the regulatory elements than the previously described RE1-9.

**Figure 2.**
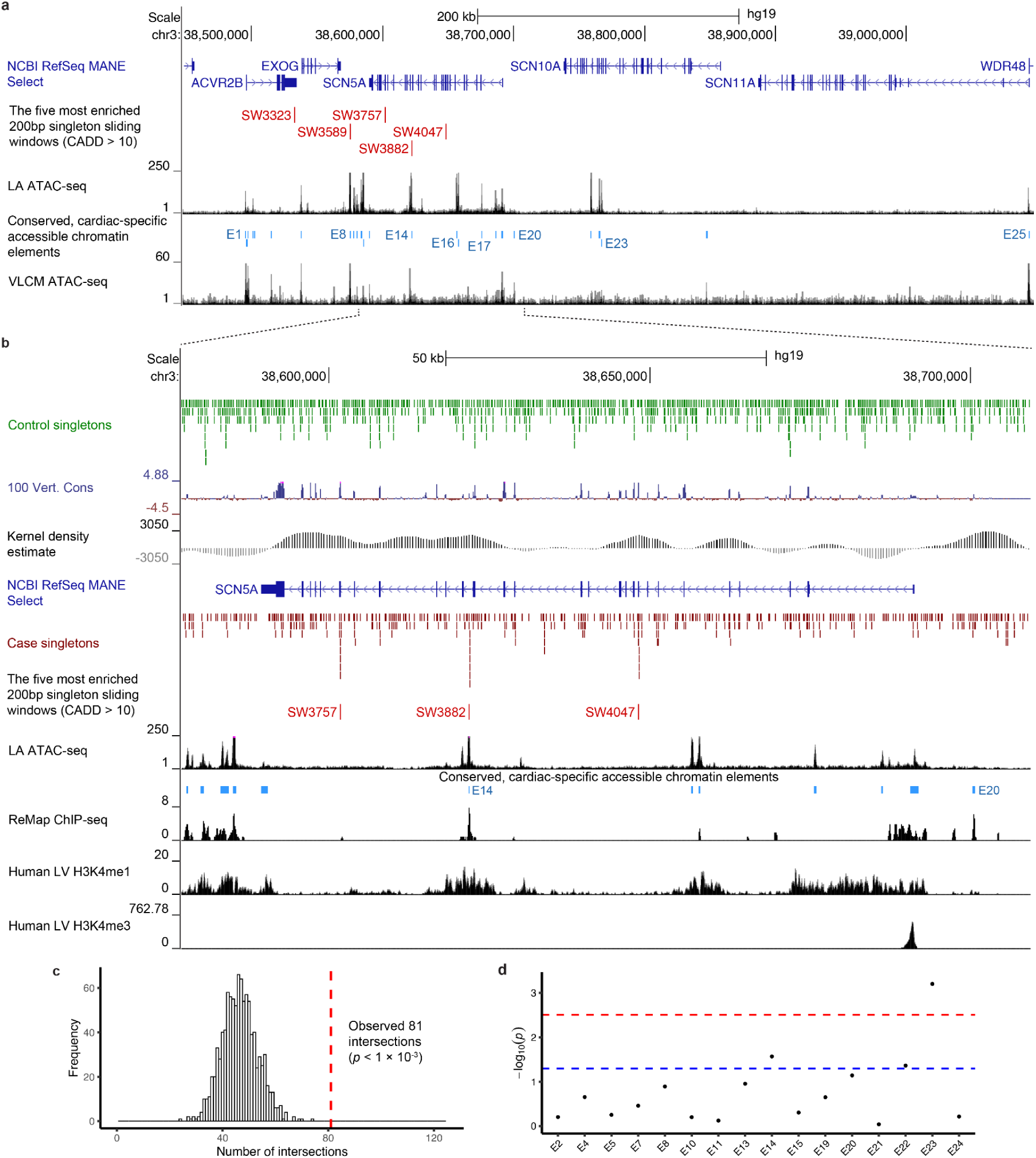
| Aggregate rare variant testing across the SCN5A TAD. (A) View of the SCN5A TAD with the open chromatin regions (left atrial ATAC-seq; ventricular-like cardiomyocyte ATAC-seq) and the candidate CREs (blue; E1-E25) shown. The most enriched sliding windows are shown in red. (B) Magnified view of the region around SCN5A. Control singleton variants are shown in green, case singleton variants are shown in red. The differential spatial density of case versus control singletons is shown by the differential kernel density estimate (grey histogram). The binding of cardiac TFs (ReMap ChIP-seq) and histone modifications associated with enhancers (human left ventricle H3K4me1) and promoters (H3K4me3) is also shown. (C) Histogram of intersections between candidate CREs and case singletons after randomization of singleton genomic positions (empirical null distribution). The dashed red line indicates the overlap count using the observed singleton positions (permutation-test p-value shown). (D) Singleton burden testing of the CREs with a cumulative minor allele count (cMAC) ≥ 5 (blue dashed line: nominal significance, α = 0.05; red dashed line: Bonferroni-corrected threshold, α_b_ = 0.05/16). LA, left atrium; LV, left ventricle; ChIP-seq, chromatin immunoprecipitation and sequencing; ATAC-seq, assay for transposase accessible chromatin and sequencing; CADD, the variant pathogenicity tool Combined Annotation Dependent Depletion.

### Rare non-coding variants in patients intersect the CREs more often than expected by chance

We asked whether BrS case singleton variants (MAC_cases_ = 1; MAC_controls_ = 0; *n* = 1871 across the region) intersected the candidate CREs in the *SCN5A* TAD (*n* = 25) more often than would be expected by chance. An *in-silico* Monte Carlo overlap enrichment experiment was performed in order to determine this (Supplemental Methods). Briefly, the case singleton variant positions were randomised (*n*_trials_ = 1000) in order to obtain the number of intersections with the candidate CREs expected by chance, then this value was compared with the observed number of intersections. There were 81 observed intersections, significantly more than the expected 46 intersections (observed/expected ratio = 1.76; *p* < 1e-3; Figure 2C).

### CRE-burden testing reveals aggregate signals at E14, E22 and E23

To identify which specific CREs were enriched, we performed burden testing of singleton variants across all CREs with a cumulative minor allele count (cMAC) ≥ 5 (16/25 met this criterion; Figure 2D). Element E23 had a Bonferroni significant enrichment of case singletons (*p* = 6.3e-4; *α*_b_ = 0.05/16); element E14 had a nominally significant enrichment of case singletons and E22 a nominally significant depletion of case singletons (*p* = 0.027 and *p* = 0.043, respectively).

### Sliding window approach highlights regions within and downstream of *SCN5A*

An unbiased, hypothesis-generating approach was also used; namely, a sliding window analysis across the *SCN5A* TAD. Briefly, 4,066 windows were defined across the region; these windows were 200 bp in length (the approximate length scale of an enhancer^39^), and each window overlapped adjacent windows by 20% on either side. Burden testing included only singleton variants with CADD > 10 and was performed for windows in which the cMAC of these qualifying variants was ≥ 5. Out of the 4,066 windows across the TAD, 15 windows met the criteria for testing, the five most enriched sliding windows are shown in red in Figure 2A. SW3323 (*p* = 0.11) and SW3589 (*p* = 0.14) overlap the 3’UTR of *ACVR2B* and CRE E8, respectively. SW3757 (*p* = 0.15), SW3882 (*p* = 0.08) and SW4047 (*p* = 0.08) overlap *SCN5A*.

We therefore examined the rare genetic variation present across *SCN5A* and its immediate flanking regions in greater detail (Figure 2B). The positions of all control singleton genetic variants (MAC_cases_ = 0; MAC_controls_ = 1; green marks) and all case singleton genetic variants (MAC_cases_ = 1; MAC_controls_ = 0; red marks) across *SCN5A* are shown, visually illustrating areas of high case/control singleton density. The differential singleton kernel density estimate (KDE), shown, aims to illustrate the difference in case-control singleton spatial density (Supplemental Methods; Figure 2B). Three major areas of high case singleton density were identified, overlapping sliding windows SW3757, SW3882 and SW4047. A concentration of case singletons in these areas can be seen, as well as peaks of high differential singleton KDE. Two of these windows, SW3757 and SW4047, overlap exons 23 and 9, respectively, of *SCN5A*, corresponding to the pore loop regions of channel domains I and III—known hotspots of pathogenic coding variants. However, SW3882 does not overlap an exon of *SCN5A*—it overlaps candidate CRE E14 inside intron 17 of *SCN5A*.

### Aggregations within *SCN5A* CRE E14 and *SCN10A* CREs E22-23

We next examined in more detail E14, prioritised by both the sliding window and CRE-burden approaches, as well as the previously mentioned CRE-burden signals at E22 and E23 (Figure 3A).

**Figure 3.**
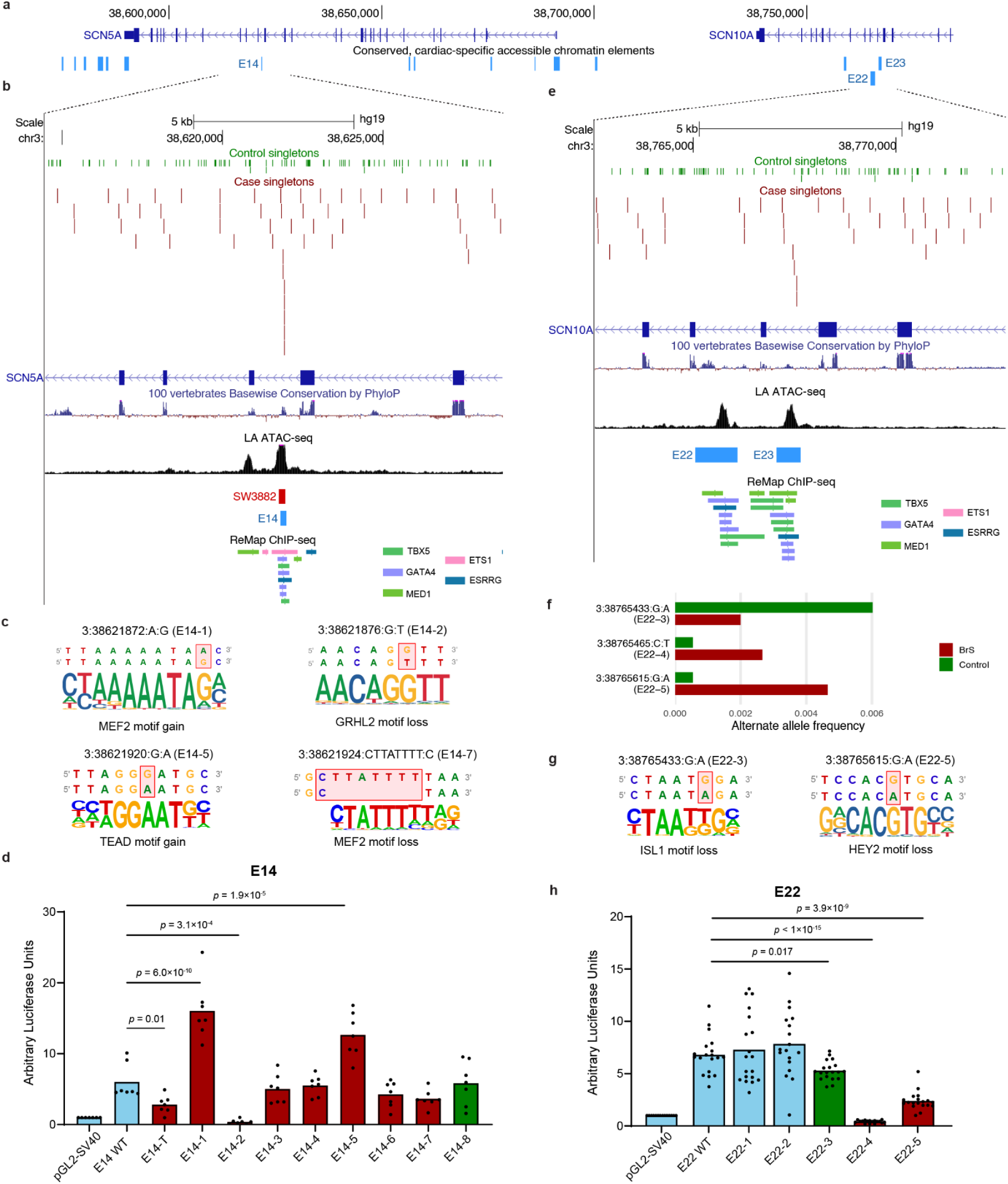
| Identification and experimental validation of non-coding variants in aggregations within SCN5A and SCN10A CREs. (A) Positions of the candidate CREs across SCN5A-SCN10A. (B), (E) Magnified views of E14 and E22/E23 respectively (blue) with positions of case/control singletons, genetic conservation (PhyloP; blue), accessible-chromatin (left atrial ATAC-seq), sliding window (red) and cardiac TF binding (ReMap ChIP-seq). (C), (G) Selected variants of interest in E14 and E22 respectively with flanking reference (top row) and alternative (bottom row) genomic sequences; affected base(s) are highlighted in red. The aligned sequence logo of the predicted affected TF motif is shown underneath. (F) The alternative allele frequency in cases and controls for low-frequency variants of interest in E22. (D), (H) Luciferase activity of constructs containing the minimal promoter (pGL2-SV40) alone or combined with wild-type or mutant E14/E22 with variants of interest. For E14, a variant previously reported in a Thai BrS cohort (E14-T; 3:38621871:A:C, GRCh37) and the E14 singletons identified in this study (case singletons: E14-1–E14-7; control singleton: E14-8). For E22, the three low frequency variants that are enriched in cases (E22-4, E22-5) or controls (E22-3) and two negative controls (E22-1, E22-2). Light blue denotes wild-type fragments, minimal promoters, or neutral variants; red, case-enriched variants; green, control-enriched variants. Statistics were performed using one-way ANOVA, followed by Dunnett’s multiple comparisons test for E14, Welch’s ANOVA followed by Dunnett’s multiple comparisons test for E17 and E22, and Kruskal-Wallis followed by Dunn’s multiple comparisons test for E23.

### An aggregation of case singleton variants within *SCN5A* intronic CRE E14

Element E14 shown in Figure 3B, overlapped by SW3882, is conserved, accessible in cardiac tissue, binds cardiac-expressed transcription factors and has flanking enhancer-associated histone modifications. Notably, a twenty-fold case-enriched low-frequency variant (3:38621871:A:C, GRCh37; referred to as E14-T) located within this same 178 bp element (which is located within the broader regulatory element, RE5, defined in prior literature^11^) has recently been described in a Thai BrS cohort^14^. This variant was found to lead to a significant reduction in *SCN5A* expression and sodium current density.

In this current work, we found 7 case singletons within the E14 CRE (E14-1 to E14-7) and 1 control singleton (E14-8); there were no other variants (for example, variants of higher allele frequency) detected in this interval. All 7 case singletons were found in samples that were negative for rare *SCN5A* coding variants. Five of the case singletons were predicted to alter TF motifs (Figure 3C; Table S6), including motifs corresponding to cardiac TFs. For instance, TEAD1 and MEF2, which have been shown to be important in cardiac development and remodelling^40–44^; and FOXP1, which has been shown to be involved in cardiac outflow tract septation^45^. Notably, one of the case singletons identified here (E14-1; Table 1) lies immediately adjacent to E14-T and is predicted to affect the same MEF2 motif. We conducted luciferase reporter assays in hiPSC-CMs on the 8 singleton variants found in this study within E14, as well as on the previously described E14-T Thai variant. In the assays we used a slightly broader CRE definition of E14, corresponding to RE5 referred to in prior literature^11^. Three case singletons significantly altered luciferase activity, in line with their computationally predicted TF motif effect (E14-1, MEF2; E14-2, GRHL2; E14-5, TEAD; Table 1; Figure 3D). The Thai E14-T variant showed the expected reduction in luciferase activity, concordant with the prior study^14^. Importantly, this reporter assay tested isolated regulatory fragments outside their native genomic context and therefore may not fully recapitulate endogenous function at the *SCN5A* locus (for instance, where such function depends on interactions with other regulatory elements, chromatin conformation or epigenetic state). Consequently, this data demonstrates that the variants qualitatively perturb E14 regulatory activity, though the direction and magnitude of their effect on endogenous *SCN5A* expression require further investigation. In the replication cohort of BrS genetic testing referrals, a significant enrichment of rare variants (gnomAD FAF_95-popmax_ < 1×10^-4^) in this CRE in cases compared to ancestry-matched controls was observed (OR = 17.0 (2.8-101.6); *p* = 0.002). Of note, as this is a referral cohort, it includes a lower proportion of definitively diagnosed BrS cases (the diagnostic yield of P/LP variants is 9.0% compared to an expected yield of ∼20% for European-ancestry BrS cases) and therefore association signals are expected to be attenuated.

**Table 1.** | Summary of prioritised non-coding variants within candidate CREs. Case singleton: AAC_cases_ = 1, AAC_controls_ = 0. Functional validation denotes a significant alteration in CRE activity in luciferase reporter assays.

| Region | Aggregate signal? | Variant (GRCh37) | Frequency | gnomAD-NFE AAF | Replication | ECG Endophenotype | TF prediction | Functional validation |
| --- | --- | --- | --- | --- | --- | --- | --- | --- |
| E14 (RE5; *SCN5A*) | Yes. Singleton CRE-burden and sliding window | 3:38621872:A:G (E14-1) | Case singleton | 0.00045 | Aggregate: OR = 17.0 (2.8-101.9) | Insufficient MAC to test in UKB | MEF2 motif strengthened | Yes |
|  |  | 3:38621876:G:T (E14-2) | Case singleton | 0 |  |  | GRHL2 motif loss | Yes |
|  |  | 3:38621920:G:A (E14-5) | Case singleton | 0 |  |  | TEAD motif gain | Yes |
|  |  | 3:38621924:CTTATTTT:C (E14-7) | Case singleton | 0 |  |  | MEF2 motif loss | Not significant |
| E17 (*SCN5A*) | No | 3:38675954:C:A (E17-4; rs186741256) | Low-frequency ($AAF_{cases} = 2.10\%$; $AAF_{controls} = 0.40\%$; OR = 5.49 (2.70-11.19)) | 0.0048 | $AAF_{cases} = 0.62\%$; $AAF_{controls} = 0.28\%$; OR = 2.26 (0.97-5.2) | QRS +1.04 ms ($p = 0.02$); PR +1.97 ms ($p = 0.01$) | HAND1 motif loss | N/A |
| E22 (RE2; *SCN10A*) | Yes. Singleton CRE-burden (control enriched) | 3:38765433:G:A (E22-3; rs187345429) | Low-frequency ($AAF_{cases} = 0.20\%$; $AAF_{controls} = 0.60\%$; OR = 0.33 (0.06-1.10)) | 0.0034 | *SCN10A* introns not present in HiC panel | PR -3.57ms ($p = 4.7e-7$); QRS $p > 0.05$ | ISL1 motif loss | Yes |
| | | 3:38765465:C:T (E22-4; rs576722503) | Low-frequency ($AAF_{cases} = 0.27\%$; $AAF_{controls} = 0.05\%$; OR = 4.87 (0.70-53.83)) | 0.00022 | | Not significant | ZNF350 motif loss | Yes |
| | | 3:38765615:G:A (E22-5; rs529833633) | Low-frequency ($AAF_{cases} = 0.47\%$; $AAF_{controls} = 0.05\%$; OR = 8.53 (1.62-84.46)) | 0.00084 | | Not significant | HEY2 motif loss | Yes |
| E23 (RE1; *SCN10A*) | Yes. Singleton CRE-burden | See Table S6 | Case singletons | See Table S6 | *SCN10A* introns not present in HiC panel | Insufficient MAC to test in UKB | See Table S6 | Insufficient baseline CRE activity |

### Low-frequency BrS-associated SNVs within *SCN10A* E22 and case singleton burden in *SCN10A* E23

As described above, singleton variant associations were observed within *SCN10A* intronic elements E22 and E23 (Figure 3E). These lie within RE2 and RE1, respectively, defined in prior literature^12^. These elements have been implicated in regulation of a short *SCN10A* transcript encoding a truncated Na_V_1.8 protein that modulates Na_V_1.5^12,46^, as well as in direct regulation of *SCN5A*^47^. In isolation, RE1 and RE2 directly drive reporter expression in mouse embryos in the sinoatrial node and precursors of the ventricular conduction system, but not in the working myocardium^12^. However, deletion of RE1 in mice reduces expression of *Scn10a* and *Scn5a* in the working myocardium, indicating that, in their native genomic context, RE1-2 may regulate *SCN10A* and *SCN5A* through their interactions with other regulatory elements in the region, potentially acting as a regulatory hub or relay^12^.

Element E22 (Figure 3E), is conserved, accessible in cardiac tissue, binds cardiac-expressed transcription factors and has flanking enhancer-associated histone modifications. E22 contained 0 case singletons and 12 control singletons; producing a nominally significant control singleton enrichment (*p* = 0.043). Other than these variants, there were also 8 low-frequency variants and 2 common variants in E22 (Table S7). Of these low-frequency variants, three variants were enriched in either cases or controls. First, a low frequency variant three-fold enriched in controls, 3:38765433:G:A (E22-3; AAF_cases_ = 0.20%, AAF_controls_ = 0.60%; OR = 0.33 (0.06-1.10); *p* = 0.07), which is predicted to lead to an ISL1 motif loss (Figure 3F,G; Table 1). Second, a low-frequency variant enriched eight-fold in cases, 3:38765615:G:A (E22-5; AAF_cases_ = 0.47%, AAF_controls_ = 0.05%; OR = 8.53 (1.62-84.46); *p* = 0.004), which is predicted to lead to the loss of a HEY2 motif (Figure 3F,G; Table 1). Third, a low-frequency variant marginally enriched in cases and predicted to disrupt a ZNF350 motif (E22-4; 3:38765465:C:T; AAF_cases_ = 0.27%, AAF_controls_ = 0.05%; OR = 4.87 (0.70-53.83); *p* = 0.06; Table 1). We explored the effect of these variants using luciferase assays, along with two negative controls—a variant found in 3 controls and no cases (E22-1; 3:38765830:T:C), and a low frequency variant with equivalent case-control frequency (E22-2; 3:38765081:T:C; AAF_cases_ = 0.07%, AAF_controls_ = 0.08%). Variants E22-3, E22-4 and E22-5 significantly altered luciferase activity; while the negative controls E22-1 and E22-2 showed no alteration (Figure 3H). E22-3, E22-4 and E22-5 were of sufficient frequency to assess their effect on ECG conduction indices in the UK Biobank (Methods; Table S8). E22-3 was significantly associated with a reduced PR interval (β = −3.57 ms; effect allele = A, *p* = 4.68×10^-7^), consistent with faster atrioventricular conduction and in line with the control enrichment observed in our BrS cohort (where conduction-favouring alleles are protective). No significant ECG associations were observed for E22-4 or E22-5. Given their lower frequencies relative to E22-3, analyses for E22-4 and E22-5 may have been underpowered. Element E23 (Figure 3E), is also conserved, accessible in cardiac tissue (ATAC-seq), binds cardiac-specific transcription factors (ChIP-seq) and has flanking enhancer-associated histone modifications. E23 harboured a significant singleton CRE-burden (*p* = 6.3e-4). The element contained 7 case singletons, 1 control singleton, 2 low-frequency variants, and 2 common variants, one of which was rs6801957, the lead SNP of the BrS GWAS^6^. Previous functional studies have implicated rs6801957 in altered RE1 (E23) activity through disruption of a T-box TF-binding motif^12,47,48^. All of the carriers of the 7 case singletons were *SCN5A* rare coding variant negative. Three of the seven case singletons possessed a sufficient MAC to be tested for ECG traits in the UK Biobank: of these, two were not associated with ECG parameters, the third (3:38767546:G:A) was associated with a prolonged QRS interval (β = +4.21 ms; effect allele = A; *p* = 5.62×10^-3^). All seven case singleton variants were computationally predicted to alter TF motifs; one of these (3:38767217:A:G) was predicted to disrupt a TF motif family with well-established roles in cardiomyocyte biology (FOXO)^49–54^. The remaining variants predominantly affected TFs with broader developmental or not clearly cardiomyocyte-specific roles. We attempted to assess all 7 case singletons, the control singleton and the common variant rs6801957 by luciferase assay.

However, the assay did not yield interpretable results because the wild-type E23 element exhibited low baseline luciferase activity relative to other CREs tested, consistent with the low activity of this isolated element in hiPSC-CMs, thereby limiting assessment of variant effects. The low activity is expected since, as described earlier, E23 (RE1) is likely to be a relay element for other CREs in the region, and not active in isolation in working cardiomyocytes^12^. We were unable to assess replication of the aggregate signals or individual variants in E22 and E23 as the BrS referral cohort gene panel does not cover *SCN10A* intronic regions.

### The identification of a large deletion of an upstream *SCN5A* CRE

Three different structural variant (SV) callers were used on the WGS dataset in an attempt to identify case-associated SVs in the *SCN5A* region (Methods). Two case singleton SVs of interest were identified in the *SCN5A* region (Figure 4A). SV1, found in one French BrS case, is a 27,750 bp heterozygous deletion of exons 6-14 of *SCN5A*, encompassing most of the first channel domain and the beginning of the second and can therefore be assumed to be pathogenic (see Figure S8 for the WGS read evidence). SV2, also found in one French BrS case, is a 10,545 bp heterozygous deletion of a non-coding region directly upstream of *SCN5A* (Figure S9). SV2, shown in Figure 4A, encompasses candidate CRE E20 and two CTCF-binding sites. SV2 was CRISPR-Cas9 engineered into hiPSC-CMs and the resulting isogenic lines were electrophysiologically characterized using automated high-throughput patch-clamp. Relative to wild-type, the mutant cells displayed a significant reduction in peak sodium current density (WT: −335.8 ± 21.2 pA/pF; SV2+/− clone 1: −251.0 ± 31.7 pA/pF; mean ± SEM; *p* = 0.0023; Figure 4B,C). There was no alteration in the voltage-dependence of activation and inactivation (Figure 4D). This pattern is consistent with reduced functional Na_V_1.5 channel abundance, as would be expected from altered *SCN5A* expression.

**Figure 4.**
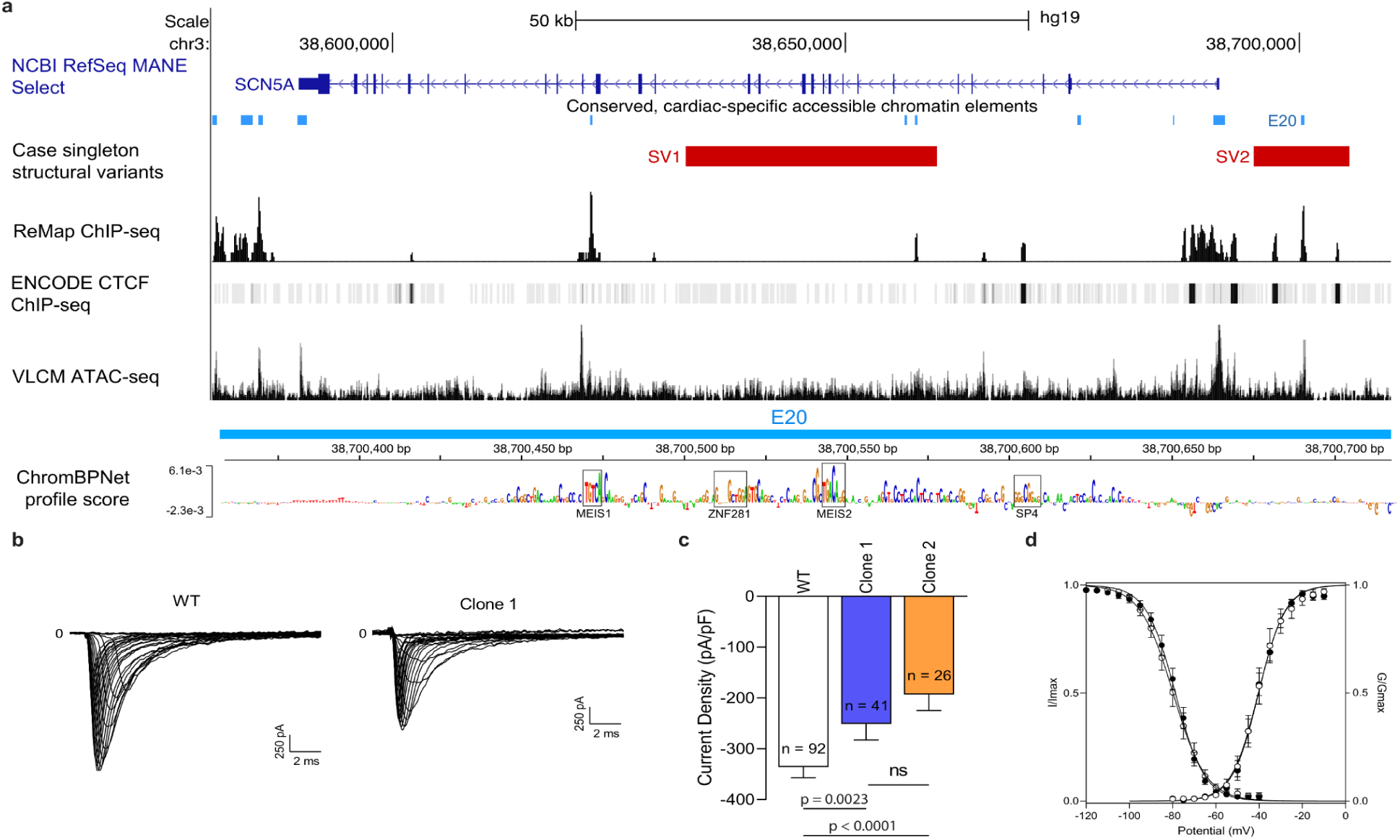
| Rare structural variation found in BrS cases in the vicinity of SCN5A. (A) View of SCN5A with candidate CREs (blue), case singleton structural variant deletions (red), cardiac TF binding (ReMap ChIP-seq), CTCF binding (ENCODE CTCF ChIP-seq) and accessible chromatin (ventricular-like cardiomyocyte ATAC-seq) shown. ChromBPNet prediction track generated from a ventricular-like cardiomyocyte training model showing enriched chromatin accessibility and cardiac TF motifs in candidate CRE E20. (B) Representative recordings of Na⁺ currents in WT hiPSC-CMs and SV2+/− clone 1 hiPSC-CMs during depolarizing steps ranging from from −80 mV to +25 mV (100 ms duration, 0.2 Hz; holding potential: −120 mV). Traces illustrate sodium current activation and inactivation in the isogenic model. (C) Quantification of peak sodium current density in WT (n = 95 cells), SV2+/− clone 1 (n = 41 cells), and SV2+/−clone 2 (n = 26 cells). Statistics were performed using the Mann-Whitney test. (D) Voltage-dependence of activation and inactivation of sodium channels in WT (n = 8 cells), SV2+/− clone 1, and SV2+/− clone 2 (n = 9 cells). SV, structural variant; VLCM, ventricular-like cardiomyocyte; WT, wild-type.

### The impact of other classes of rare non-coding variation

We explored if other forms of rare and low-frequency non-coding variation within the *SCN5A* region, that do not belong to the CRE-variant category, were associated with BrS, namely, variants that disrupt CTCF-binding sites, that disrupt miRNA/RBP-binding in UTRs and those that fall within exons of ncRNAs in the region.

### Limited evidence of CTCF-disrupting SNVs and indels

CTCF-bound sites help organize the three-dimensional chromatin topology of this region by constraining cohesin-mediated loop extrusion, thereby establishing insulating TAD boundaries and intra-domain chromatin loops that help bring CREs and promoters into close physical proximity. Variants that weaken CTCF binding could therefore alter *SCN5A* expression by reducing normal CRE-promoter contact probabilities or by permitting ectopic regulatory interactions across compromised TAD boundaries. We sought to identify case-associated genetic variants which disrupt the binding of CTCF proteins in any of the candidate CTCF-binding regions, C1-C18, in the *SCN5A* region. Aggregate variant testing of all rare and low-frequency variants (AAF_gnomAD-NFE_ < 1%) in each of the 18 candidate CTCF-binding regions was performed (Supplemental Methods). No candidate CTCF-binding region reached Bonferroni-corrected significance (Figure S7B). Two regions reached nominal significance; CTCF-binding region C7 (SKAT *p* = 0.03), located directly upstream of *SCN5A* and CTCF-binding region C14 (burden *p* = 0.02), located inside intron 2 of *SCN11A.* We assessed the predicted TF motif impact for all variants within these two regions. Only one was predicted to affect a CTCF-motif—a case singleton variant, 3:38692882:T:C, inside C7 which was predicted to weaken a CTCF-binding motif (the variant is absent from gnomAD and the UKB; carrier is *SCN5A* rare coding variant negative; Figure S7C).

### No significant *SCN5A* UTR burden was detected

We tested for case enrichment of variants (AAF_gnomAD-NFE_ < 1%) in the UTRs (combined 3’ and 5’) of *SCN5A* and *SCN10A*. The *SCN10A* UTR mask did not fulfill the minimum cMAC ≥ 5 for inclusion; the *SCN5A* UTR mask did, but showed no significant enrichment (Figure S7D). The 41 singleton and low-frequency variants (AAF_gnomAD-NFE_ < 1%) in the *SCN5A* 3’UTR were annotated using the *in-silico* tool RegVar^21^. Several variants were annotated as overlapping predicted miRNA target sites and/or as altering RBP-binding motifs, however there was no clear enrichment of these variants among case singletons or case-enriched low-frequency variants. Taken together, the data here does not support a substantial contribution of rare *SCN5A* UTR variants to BrS susceptibility.

### No evidence of burden in ncRNA genes near *SCN5A*

Some non-coding RNAs act in *cis* to regulate neighbouring genes through mechanisms including antisense interactions, transcriptional interference, or chromatin modulation. Rare variants within exonic regions of ncRNAs proximal to *SCN5A* may plausibly influence its expression, hence we burden tested these regions (AAF_gnomAD-NFE_ < 1%; Supplemental Methods). There are 5 ncRNA genes in the *SCN5A* TAD, only one ncRNA gene reached the minimum cMAC ≥ 5 for inclusion: *ACVR2B-AS1*. It showed no significant burden (Figure S7E).

### WGS reveals low-frequency non-coding and structural variation underlying Brugada syndrome GWAS SNPs

Unlike SNP-array genotyping, WGS enables the interrogation of variants across all classes, including structural variation, and across all allele-frequencies. The largest to date European-ancestry BrS GWAS^6^ identified eight conditionally-independent genome-wide significant common SNPs at the *SCN5A*-*SCN10A* locus. These SNPs may not necessarily be functional or causal; instead, they could be in linkage disequilibrium (LD) with the causal variant(s). Using WGS, we aimed to identify putatively causal non-coding variation that could underlie, at least in part, some of the BrS GWAS signals, exploring a wider spectrum of allele frequencies and variant classes than those captured by the GWAS. To this end, we integrated epigenomic data with a low-frequency single-variant analysis, followed by conditional analyses. In addition, our SV calling identified a common SV upon which we performed conditional analyses.

### Case-enriched low-frequency SNV within a CRE in intron 1 of *SCN5A*

A single-variant analysis across the *SCN5A* TAD (chr3:38488336-39139033) with an *AAF*_gnomAD-popmax_ < 1% filter identified one Bonferroni-corrected significant low-frequency variant (*p* = 8.9e-07), that would not have been considered by the BrS GWAS (Figure 5A-B). This five-fold case-enriched variant, 3:38675954:C:A (*AAF*_cases_ = 2.1%, *AAF*_controls_ = 0.4%, OR = 5.49 (2.70-11.19)), is located inside candidate CRE E17 in the downstream region of intron 1 of *SCN5A*, and is predicted to lead to the loss of a HAND1 TF motif (Figure 5A and 5C; Table 1). HAND1 has an established role in the development of the ventricular conduction system^55^. Conditioning on this SNV in a single-variant analysis (*SCN5A* TAD; all frequencies) reduced the significance of one of the BrS GWAS SNPs: rs41310232 (Figure 5D). The low-frequency SNV 3:38675954:C:A is in complete linkage disequilibrium (*D*′ = 1) with the common SNP rs41310232, albeit with a low *r*² (0.06), reflecting their large difference in allele frequency. The GWAS SNP rs41310232 is not located within a candidate CRE. We hypothesize that rs41310232 tags the putatively functional low-frequency SNV 3:38675954:C:A in E17. Association testing of this SNV in the independent Health in Code BrS referral cohort uncovered a higher frequency of the effect allele in cases compared to controls (*AAF*_cases_ = 0.62%; *AAF*_controls_ = 0.28%; OR = 2.26 (0.97-5.2); p = 0.061; Table S10), with the effect size attenuated as expected by the referral nature of this cohort. In addition, in the UKB, it was found to be associated with prolonged QRS and PR intervals (+1.04 ms QRS, *p* = 0.02; +1.97 ms PR, *p* = 0.01; Table S8). However, this SNV only accounts for part of the SNP signal, suggesting the rs41310232 SNP could be tagging additional low-frequency functional variants. Conditioning on the putatively functional 3:38675954:C:A SNV in the low-frequency single-variant analysis (*AAF*_gnomAD-popmax_ < 1%) revealed a new lead variant (Figure S11B): a second intron 1 low-frequency SNV, 3:38675003:T:G (*AAF*_cases_ = 1.4%; *AAF*_controls_ = 0.38%; OR = 4.74 (2.28-9.86); *p* = 2.41e-05; rs45527135). This variant also replicated in the Health in Code cohort (*AAF*_cases_ = 0.54%; *AAF*_controls_ = 0.23%; OR = 2.38 (1.48-3.85); *p* = 0.0011; Table S10). The low-frequency SV 3:38675003:T:G is also in complete LD with the common SNP rs41310232 (*D’* = 1) and further reduces the significance of rs41310232 upon conditioning (Fig S13C), suggesting it could be a second putatively functional low-frequency SNV tagged by the SNP.

**Figure 5.**
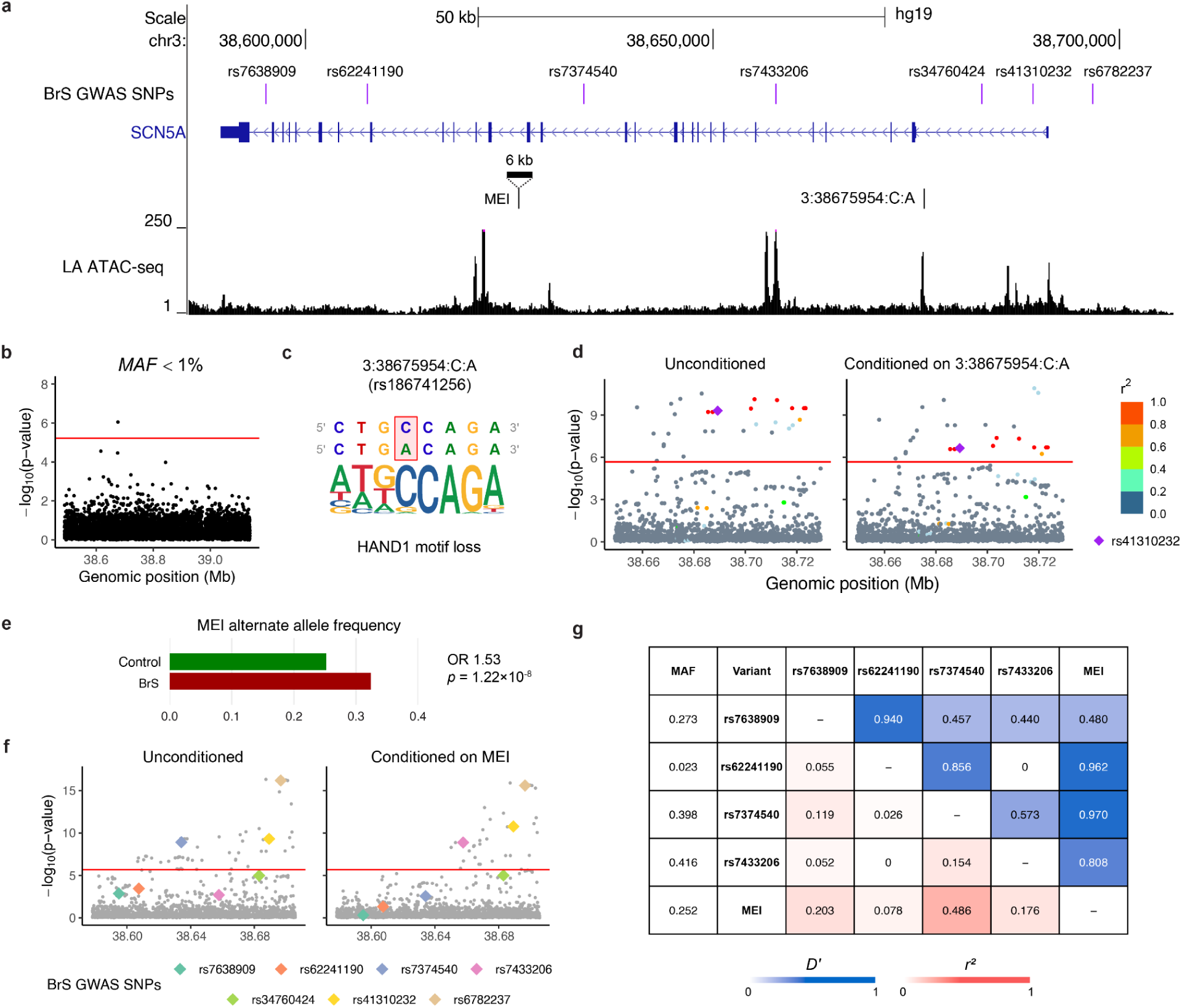
| Putatively functional low-frequency SNV and common mobile element insertion underlying BrS GWAS SNPs. (A) View of SCN5A with 7 of the 8 conditionally-independent BrS GWAS SNPs (purple) at the SCN5A-SCN10A locus from Barc et al. (2022) (the 8th is in SCN10A, not shown here). The mobile element insertion (MEI; black, with 6 kb insertion sequence), the low-frequency SNV (3:38675954:C:A; black) and accessible-chromatin (left atrial ATAC-seq) are shown underneath. (B) Single-variant association analysis across the SCN5A region (AAF_gnomAD-popmax_ < 1%; Bonferroni-corrected threshold in red). (C) The low-frequency SNV (3:38675954:C:A) with flanking reference (top row) and alternative (bottom row) genomic sequences; affected base is highlighted in red. The aligned sequence logo of the predicted affected TF motif is shown underneath. (D) Single-variant association analysis across the SCN5A region (all AAF; Bonferroni-corrected threshold in red) unconditioned (left) and conditioned on the low-frequency SNV, 3:38675954:C:A (right). The colours indicate the level of LD (r^2^) of the variants with the BrS GWAS SNP, rs41310232 (purple diamond). (E) Mobile-element insertion alternate allele frequency in cases/controls (OR and p-value from logistic regression). (F) Single-variant association analysis across the SCN5A region (all AAF; Bonferroni-corrected threshold in red) unconditioned (left) and conditioned (right) on the MEI. The 7 coloured diamonds indicate the 7 BrS GWAS SNPs in this region. (G) Linkage disequilibrium heatmap (r^2^ in red, D’ in blue) for the MEI and the 4 BrS GWAS SNPs affected by MEI conditioning. MEI, mobile element insertion; MAF, minor allele frequency.

### A common mobile element insertion underlying several BrS GWAS SNPs

A common mobile element insertion (MEI) was detected in intron 16 of *SCN5A* (3:38626065:C:<INS>; Figure 5A, Figure S10). The allele frequency of the MEI was higher in cases than controls (32.4% vs 25.2%; OR = 1.53 (1.32–1.76); *p* = 1.22e-8; Figure 5E). The MEI is a 6,018 bp LINE1 retrotransposon insertion, for which standard SV callers showed limited sensitivity, necessitating the use of MELT^32^, a tool specifically designed to detect mobile element insertions. In a single-variant analysis (Figure 5F), conditioning upon the MEI substantially reduced the significance of 3 of the 7 BrS GWAS SNPs at the *SCN5A* locus (rs7638909, rs62241190 and rs7374540) and increased the significance of one other (rs7433206). The MEI is in LD with these 4 SNPs (Figure 5G), especially with rs7374540 (*r*^2^ = 0.486/D’ = 0.970) which is not located within a candidate CRE and has no other strong causal candidates in LD (*r*^2^ > 0.1) with it. We hypothesize that this 6 kb intronic insertion has a higher prior probability of causality than a single-nucleotide polymorphism in a non-CRE region, for example by perturbing 3D chromatin topology or introducing novel transcription factor binding sites. We therefore propose that these GWAS SNPs may tag this putatively functional retrotransposon insertion, identified via WGS.

## Discussion

### The complex genetic architecture of BrS at the *SCN5A* locus

The *SCN5A* region is central to the genetics of BrS. Rare loss-of-function coding variants in *SCN5A* are established primary causes of the disease, while common risk variants within the locus have been identified in GWAS of BrS and associated with ECG indices of slower cardiac conduction, consistent with reduced conduction reserve. However, the full spectrum of variation at this locus, and the mechanisms by which it regulates *SCN5A* expression and function, remain incompletely understood—especially with regard to rare non-coding variation. This study systematically interrogated all forms of genetic variation at this locus, with an emphasis on rare non-coding variation, using the largest cohort to date of whole genome sequenced BrS patients. Our findings suggest that this region harbours a complex genetic architecture in which variants across a broad range of allele frequencies and variant classes influence *SCN5A* expression and confer risk for BrS.

We identified CRE-altering variants of differing population frequency at both *SCN5A* and *SCN10A* that are robustly associated with BrS. Significant aggregations of rare singleton non-coding variants were observed for three cardiac CREs. In particular, a robust enrichment signal was detected within a conserved enhancer (E14; referred to as RE5 in prior literature^11^) located in intron 17 of *SCN5A*, harbouring several variants identified in our cohort that were predicted to disrupt cardiac TF binding motifs and shown to alter transcriptional activity in reporter assays. The importance of this element to BrS pathophysiology is also highlighted by a functionally validated rare variant that occurs in 4% of BrS cases in Thailand, contributing to the increased disease prevalence in this region^14^. A similar enrichment of rare singleton non-coding variants was observed in an intronic enhancer of *SCN10A* (E23/RE1), which harbours the lead SNP from the BrS GWAS^6^ and has been experimentally shown to regulate *SCN5A* as well as a short *SCN10A* isoform that encodes a truncated Na_V_1.8 protein that modulates Na_V_1.5 activity^12^.

We also identified and functionally validated several low-frequency regulatory variants that conferred BrS risk, with effect sizes intermediate between rare pathogenic variants and GWAS SNPs. Among those with the strongest evidence was a five-fold case-enriched variant located within E17 in intron 1 of *SCN5A* (3:38675954:C:A; Table 1) which was associated with BrS risk in both discovery and replication cohorts. In the UK Biobank, the variant influenced PR and QRS intervals in the expected direction (with the BrS risk allele being associated with indices of slower conduction). The E17 region has previously been shown in a mouse model to repress *SCN5A* expression^56,57^ and harbours rare functionally validated variants in atrial fibrillation patients^58^. Additionally, three low frequency variants were identified within an intronic enhancer (E22) of *SCN10A*, including a variant enriched eight-fold in cases and predicted to disrupt a HEY2 motif; notably *HEY2* is the strongest BrS GWAS locus outside the *SCN5A–SCN10A* region^6,59^. Another low frequency variant, altering an ISL1 motif, was three-fold control-enriched and concordantly associated with faster conduction on the ECG—a putatively protective variant.

WGS also revealed that part of the common variant association signal at this locus may be driven by variation not captured by SNP arrays. Specifically, a LINE-1 MEI within intron 16 of *SCN5A* was enriched in cases and appeared to account for a substantial proportion of the association signal observed at multiple GWAS SNPs through linkage disequilibrium. MEIs may influence gene regulation through multiple mechanisms, including perturbation of local *cis*-regulatory activity, introduction of novel TF-binding sites, alteration of chromatin looping, and epigenomic repression via local histone modifications that recruit heterochromatin forming factors^60–63^. Dilated cardiomyopathy caused by a LINE-1 MEI at the *TTN* gene has been previously described in the literature^64^. The observation that such an insertion may underlie part of the GWAS signal illustrates how GWAS signals may sometimes represent proxies for underlying structural or low-frequency variants, emphasizing the value of WGS for resolving the genetic basis of complex loci and suggesting that structural variants may contribute more broadly to disease-associated loci identified through GWAS.

Other structural variants affecting local regulatory connections were also detected via WGS. In particular, a 10.5 kb deletion encompassing an upstream CRE and CTCF-binding sites was identified in one BrS case and, when introduced into hiPSC-CMs using CRISPR-Cas9, resulted in reduced peak sodium current density. These findings demonstrate how structural variation can alter local chromatin organization and disrupt regulatory interactions that modulate *SCN5A* expression, providing an additional mechanism by which sodium channel expression may be altered in BrS.

Beyond these primarily CRE-disrupting variants, we found limited evidence that other classes of non-coding variation contribute substantially to BrS at this locus. Rare SNVs disrupting CTCF-binding sites showed only nominal signals, and no burden was detected in the nearby ncRNA gene *ACVR2B-AS1*. Similarly, we did not observe deleterious deep-intronic splice variants in *SCN5A*, nor a coding burden in *SCN10A*/*SCN10A-short*, nor an *SCN5A* UTR burden. These findings therefore help refine the spectrum of variant classes most likely to contribute to disease susceptibility in this region, suggesting that CRE-altering variation may be the most important non-coding variant class at this locus for BrS.

### The importance of a targeted, integrative analysis

Our results illustrate the methodological challenges associated with identifying rare and low frequency disease-associated non-coding variants in rare conditions. Since rare non-coding variants occur as singletons or very low-frequency variants, there is limited statistical power for single-variant association testing, which necessitates aggregate approaches. However, defining appropriate regions for burden testing within the vast and still poorly understood non-coding genome remains challenging. Small testing regions can result in very low cumulative minor allele counts and a substantial multiple-testing burden, while larger regions dilute signals with neutral variation. As a result, unbiased genome-wide non-coding rare-variant association studies may have limited power to detect signals, particularly in modestly sized cohorts typical of rare diseases and where WGS remains costly. These difficulties are especially evident in sliding-window analyses, which involve very large numbers of tests—notably, several recent large non-coding rare-variant studies reported no genome-wide significant windows^65,66^. There is currently no standard analytical framework for rare non-coding association studies.

In this study, we addressed the above challenges through a targeted and integrative analytical strategy. By focusing on the 600 kb topologically associating domain encompassing the principal disease gene in BrS, *SCN5A*, and incorporating cardiac-specific epigenomic datasets to define candidate regulatory elements, we restricted the testing space to biologically plausible regions. This targeted approach both reduced the multiple-testing burden and increased statistical power. Several signals identified in this study, including the enhancer within intron 17 of *SCN5A*, would likely have been missed by genome-wide approaches alone.

This underscores the importance of carefully and contextually incorporating locus-specific biological priors into rare non-coding association analyses. In contrast to the relatively coarse annotations often applied in genome-wide scans, we integrated cardiac-specific epigenomic datasets to delineate candidate CREs at high resolution across the *SCN5A* locus. More broadly, successful non-coding variant discovery in rare disorders will likely require careful integrated strategies that combine WGS with detailed tissue-specific functional genomics and appropriate experimental validation. Incorporating other lines of supporting orthogonal evidence will also be important, such as relevant endophenotype associations in population-scale biobank data and replication in independent disease cohorts. For conditions such as BrS, where a single gene plays a central role but much of the heritability remains unexplained, focusing on the regulatory landscape surrounding known disease genes may provide a particularly powerful approach.

Future studies could further refine our understanding of the regulatory architecture of the *SCN5A* locus by integrating WGS with high-resolution chromatin conformation mapping, enabling direct assessment of how WGS-detected variants, such as structural deletions and retrotransposon insertions, alter the three-dimensional chromatin topology of the region. More broadly, the decreasing cost of WGS has led to increasingly large genomics datasets, for instance those generated by large biobank initiatives such as the UK Biobank. Combined with advances in AI-based models^67,68^ that integrate epigenomic data to prioritise regulatory variants and regions with high fidelity, alongside high-throughput functional assays including massively parallel reporter assays (MPRAs)^69,70^, these developments may enable more effective genome-wide rare non-coding variant association studies. Continued improvements in statistical methodology for rare variant analysis, such as dynamic-sliding window frameworks, may further enhance the power of such analyses^71,72^.

### Clinical relevance and implications for polygenic risk scores

Clinical genetic testing for BrS currently focuses largely on rare coding variants in *SCN5A*. Although our findings indicate that rare non-coding SNVs and non-coding structural variants at the same locus may also contribute to disease susceptibility, more efforts may be needed before they are incorporated into routine clinical testing. The clinical interpretation of non-coding variants remains challenging. Although rare non-coding variants can be assessed using adaptations of the ACMG/AMP evidence framework^73^, the evidence lines typically used to establish pathogenicity for *SCN5A* coding variants translate poorly to the non-coding setting. Protein hotspot domain^4^ evidence has no direct analogue, variant enrichment evidence in large-scale datasets is typically unavailable, computational predictors are underdeveloped and complex non-coding variant validation experiments are required rather than the established functional assays in heterologous cell lines^74^. In this study, population-scale biobank data enabled the evaluation of certain variants with respect to ECG traits such as PR and QRS intervals, providing orthogonal evidence supporting their biological relevance and connecting these variants to clinical phenotypes. Nevertheless, clinical translation will require more extensive experimental validation, replication in independent BrS cohorts, and analyses in larger WGS datasets. Although the diagnostic yield of putatively causal rare variants identified here was modest, our findings highlight the importance of low-frequency variants with intermediate effect sizes. While these variants are unlikely to represent single causes of disease, their effects are substantially larger than those of typical GWAS SNPs and they may contribute meaningfully to disease susceptibility. Consideration of such intermediate-effect variants in future clinical genetics workflows may therefore be important^75,76^.

Our findings also have implications for the construction and interpretation of polygenic scores (PGS). The observation that part of the GWAS signal appears to be accounted for by an MEI highlights how GWAS SNPs may act as proxies for underlying undetected functional biological variation. While such proxy relationships enable SNP-based PGS to statistically capture part of the disease-associated genetic variation, they may not fully represent contributions from low-frequency variants or structural variants which are often incompletely tagged. As WGS becomes increasingly accessible, incorporating a broader spectrum of genetic variation, including low-frequency and structural variants, into genetic risk models may further refine risk prediction beyond conventional SNP-based PGS. For BrS, integrating these additional genetic contributors into polygenic and clinical risk models could improve future risk stratification.

### Limitations

Some limitations should be considered. First, although our cohort represents a relatively large WGS cohort for a rare disease and the largest BrS WGS cohort assembled to date, statistical power remains limited for detecting very rare regulatory variants. Larger international WGS datasets will be required to systematically characterize the contribution of rare non-coding variation to BrS. Second, replication of rare non-coding variant associations remains challenging in rare diseases due to the limited availability of independent WGS cohorts of sufficient size. Finally, while experimental systems such as luciferase reporter assays provide useful functional evidence, they represent simplified systems that cannot fully recapitulate the endogenous chromatin context, long-range regulatory interactions, and three-dimensional chromatin topology of the intact human heart. In addition, these experiments remain labour-intensive and are difficult to apply systematically to large numbers of variants. Similarly, motif-disruption predictions provide useful hypotheses but are not definitive evidence of regulatory effects.

### Conclusions

In conclusion, analysing the largest whole genome sequenced cohort of BrS patients to date, we used a targeted, integrative approach focused on the 600 kb *SCN5A* TAD to identify disease-associated rare non-coding variants. This approach emphasised the careful integration of tissue-specific functional genomic data with WGS at a known disease gene locus, supported by evidence from reporter assays, population-scale biobank data, and independent cohort replication. Such a strategy may prove valuable for other rare disorders where modest sample sizes limit the power of non-coding rare-variant studies. Among our findings are rare and low frequency CRE-disrupting variants associated with BrS risk, a 10.5 kb CRE-deletion upstream of *SCN5A* and a common retrotransposon insertion that appears to underlie part of the GWAS signal at this locus. Collectively, these observations highlight the complexity of the genetic architecture of BrS at this locus and represent a step forward in explaining a proportion of the unresolved genetic basis of BrS using whole genome sequencing.

## Data and code availability

The code and summary-level data from this study is available upon reasonable request from the corresponding author.

## Acknowledgements

This study was supported by the Agence Nationale de la Recherche (ANR-14-CE10-0001-01 to R.R.), France Génomique (ANR-10-INBS-009, ‘Brugada-seq’ to R.R.) and the Fondation pour la Recherche Médicale (Equipe FRM DEQ20140329545 to J.J.S.). We thank the biological resource centre for biobanking (CHU Nantes, Nantes Université, Centre de ressources biologiques (BB-0033-00040), F-44000 Nantes, France). We are most grateful to the Genomics Core Facility GenoA, member of Biogenouest and France Genomique and to the Bioinformatics Core Facility BiRD, member of Biogenouest and Institut Français de Bioinformatique (IFB) (ANR-11-INBS-0013) for the use of their resources and their technical support. We are grateful to the support of the GenOmic variability in heaLth & Disease “GOLD”, a national transversal research project supported by INSERM. M.B. was supported by IRP-VERACITIES—New Mechanisms for VEntricular ARrhythmia and CardIomeTabolic DIseasES, an I-SITE NExT health and engineering initiative (Ecole Centrale & Nantes University), by the IRP—GAINES—Genetic Architecture IN cardiovascular disEaSes funded by INSERM and CNRS and “Fondation pour la Recherche Médicale” (FRM: FDT202204014986). E.R.B and Y.D.W acknowledge funding from the Robert Lancaster Memorial Fund, the National Institute of Health Research and the Higher Education Funding Council for England. R.W. was supported by the Academy of Medical Sciences (grant SBF0011\1180). A.M.G and D.M.R are supported by R01HL164675; A.M.G is supported by R00 HG010904. S.J.J. was supported by a doctoral fellowship from Amsterdam UMC. This work was supported by grants from the Dutch Heart Foundation (PREDICT and PREDICT2 to C.R.B and A.A.M.W; 03-007-2022-0035 to S.J.J., and 03-003-2021-T061 to P.G.P.), the Dutch Research Council (VICI award to C.R.B.), the KIC program through the Dutch Research Council and Dutch Heart Foundation (PRECISE project to C.R.B. and S.J.J.), and the European Research Council (Horizon Pathfinder Challenges 01 – grant 101115295 – NaV1.5 to J.B., M.G. and C.R.B.; DCM-NEXT project to C.R.B.). The authors thank all the participants and professionals contributing to the UK Biobank. All analyses for this study were conducted under UK Biobank application number 176602.

## Declaration of interests

J.H.V. reports to have sponsored research agreements with Biogen, Eli Lilly, Trace and Astra Zeneca. The other authors declare no competing interests.

## Supplemental information

Supplemental information can be found online.

## Notes

### Author Declarations

The Institutional Medical Ethics Committee of Nantes University Hospital gave ethical approval for this work (reference DC-2011-1399). The Medical Ethics Review Committee of Amsterdam University Medical Center gave ethical approval for this work (reference W20_226 #20.260). The West London Research Ethics Committee of the UK Health Research Authority gave ethical approval for the St Georges University Hospitals NHS Foundation Trust component of this work (REC reference 10/H0803/121; IRAS project ID 53663). The London-Bromley Research Ethics Committee of the UK Health Research Authority gave ethical approval for the University College London component of this work (REC reference 15/LO/0549). All participants provided written informed consent.

