## Supplemental Methods and Figures for "Whole-genome sequencing implicates rare, low-frequency and structural non-coding variation at the *SCN5A* locus in Brugada syndrome"

### Contents

**Supplemental Methods**
- Whole-genome sequencing pipeline
  - Upstream pipeline
  - Downstream pipeline
- Variant annotation and regions of interest
  - SpliceAI prioritisation criteria
  - Epigenomic datasets and defining regions of interest
  - Transcription factor motif disruption prediction
  - ChromBPNet prediction
- Genomic analyses
  - Cis-regulatory element and CTCF-binding site aggregate testing
  - Sliding window aggregate testing
  - Single-variant analysis
  - Monte-Carlo overlap enrichment experiments
  - Structural variant calling and mobile element insertion conditional analysis
  - Differential singleton kernel density estimation
  - Gene-centric aggregate testing
  - Replication of associations: Health in Code genetic analysis
  - Effect on ECG endophenotypes in the UK Biobank
- Cis-regulatory element functional validation
  - Generation and preparation of hiPSC-CMs for transfection experiments
  - Cell culture and transfection luciferase assays
  - Fragment coordinates (hg38)
- Structural variant functional validation
  - Patient recruitment and deletion identification
  - Generation of hiPSCs and ventricular-like differentiation
  - Generation of the isogenic model containing the 10 kb deletion on hiPSCs
  - Transcriptomic analyses of the isogenic hiPSC-CM-V model
  - Electrophysiological studies
- Splice variant functional validation
  - Minigene assays
  - CRISPR-Cas9 iPSC experiments

**Supplemental Figures**
- Figure S1: Variant QC metrics
- Figure S2: Heterozygosity and missingness sample QC
- Figure S3: Sample ancestry (global)
- Figure S4: Sample ancestry (European)
- Figure S5: Case-control ancestral PCs

Figure S6: Functional studies of potential splice-altering variants
Figure S7: CTCF, UTR and exonic ncRNA genetic variation
Figure S8: Read evidence for SV1
Figure S9: Read evidence for SV2
Figure S10: Read evidence for the mobile element insertion
Figure S11: Low-frequency single-variant conditional analysis
**Supplementary References**

### Supplemental Methods

#### Whole-genome sequencing pipeline

##### Upstream pipeline

Peripheral blood samples were collected from the cases and controls, from which DNA was extracted. This was whole-genome sequenced by the Illumina FastTrack service on HiSeqX with a PCR-free library. The coverage was 30x. Raw FASTQ sequence reads were aligned to the human reference genome, build GRCh37, using BWA-MEM 0.7.1247[1]. These SAM files were converted to BAM files using SAMtools 1.3.148[2]. The data from the alignment and MarkIlluminaAdapters was merged with MergeBamAlignment, this was then sorted with Sambamba 0.6.649[3]. In the case that a sample belonged to multiple read groups, the read groups were merged with sambamba merge. The BAM files were split into chromosomes and cleaned per chromosome using Picard CleanSam; duplicates were removed with Sambamba markdup. The indels were realigned using GATK 3.5 RealignerTargetCreator and IndelRealigner. Then, all of the chromosomes and non-chromosomes were merged back into one BAM file using sambamba merge. Using GATK HaplotypeCaller 50[4], variants were joint-called across the UK, Dutch and French BAMs (n = 2652), only in the region chr3:38000000-39500000, covering the *SCN5A* gene and neighbouring genes. All of the gVCFs were merged utilizing GATK3.8 CombineGVCFs, these were called using GenotypeGVCFs. The SNPs and indels were recalibrated using GATK VariantRecalibrator and ApplyRecalibration. Using SNPsift/SNPeff 4.1, the variants were annotated. Snakemake[5] was used to implement the pipeline. The resultant pVCF was passed through the variant QC pipeline detailed below, leaving 27336 variants.

- Exclusions of regions of the genome (in the Makefile)
    - Regions excluded from ENCODE file (low mappability): https://hgdownload.cse.ucsc.edu/goldenPath/hg19/encodeDCC/wgEncodeMapability/wgEncodeDukeMapabilityRegionsExcludable.bed.gz The Duke Uniqueness tracks display how unique each sequence is on the positive strand starting at a particular base and of a particular length.
    - Non-sequenced regions of the genome (i.e. centromere, telomere) excluded : https://hgdownload.cse.ucsc.edu/goldenPath/hg19/database/gap.txt.gz
    - HengLi regions excluded : https://doi.org/10.1093/bioinformatics/btu356 > "erroneous realignment in low-complexity regions and the incomplete reference genome with respect to the sample as the two major sources of errors"
- Exclusions of variants (in the snakemake file)
    - exclusion of variant with more than 1 ALT (keep only bi-allelic variants)
    - exclusion of variants with a genotyping rate < 95%
    - exclusion of variants with at least one homozygous alt (HOMVAR > 0) and 0 heterozygous

- exclusion of variants with an allele count = 0 (remaining variants after individual exclusion)
- exclusion of spanning deletion
- exclusion of variants with average depth < 10 or average depth > 100
- exclusion of variants with genotyping quality < 90
- exclusion of singleton (allele balance between 0.2 and 0.8)
- exclusion of variants with DukeMapability != 1 (wgEncodeDukeMapabilityUniqueness35bp.bigWig)
- exclusion of variants in gnomAD genome with flag :
    - gnomAD genome AC0 > ##FILTER=<ID=AC0,Description="Allele count is zero after filtering out low-confidence genotypes (GQ < 20; DP < 10; and AB < 0.2 for het calls)">
    - gnomAD genome inbreeding coefficient > ##FILTER=<ID=InbreedingCoeff,Description="InbreedingCoeff < -0.3"> Excess heterozygotes defined by an inbreeding coefficient < -0.3
    - gnomAD genome random forest > ##FILTER=<ID=RF,Description="Failed random forest filtering thresholds of 0.2634762834546574, 0.22213813189901457 (probabilities of being a true positive variant) for SNPs, indels">
- exclusion of variants in gnomAD exome with flag :
    - gnomAD exome AC0 > ##FILTER=<ID=AC0,Description="Allele count is zero after filtering out low-confidence genotypes (GQ < 20; DP < 10; and AB < 0.2 for het calls)">
    - gnomAD exome inbreeding coefficient > ##FILTER=<ID=InbreedingCoeff,Description="InbreedingCoeff < -0.3"> Excess heterozygotes defined by an inbreeding coefficient < -0.3
    - gnomAD exome random forest > ##FILTER=<ID=RF,Description="Failed random forest filtering thresholds of 0.2634762834546574, 0.22213813189901457 (probabilities of being a true positive variant) for SNPs, indels">
- exclusion of variants with hardy-weinberg p-value > 1e-15 (autosomes)
- exclusion of variants filtered in gnomAD hg38 (AC0 / AS_VQSR / InbreedingCoeff)

#### Downstream pipeline

After this, VCFtools[6], BCFtools[2] and PLINK[7] were used for further, downstream, quality control. SNVs with the flag ‘VQSRTrancheSNP99.00to99.90’ or ‘VQSRTrancheSNP99.90to100.00’ were removed. Indels with the flag ‘VQSRTrancheINDEL99.00to99.90’ or ‘VQSRTrancheINDEL99.90to100.00’ were removed. This left 25198 variants. Variants with a Phred quality score < 60 were removed, leaving 25162 variants. Variant QC metrics can be found in Figure S1. UK and Dutch samples (pre-QC: n = 424 cases, n = 801 controls) failing sample QC, determined during a prior project, were removed. Specifically, UK/Dutch samples were excluded if they had a heterozygosity rate > 0.4 (n = 8 controls), had an inbreeding F > 0.1

(n = 2 controls), were closely related (PI_HAT > 0.1; n = 7 cases, n = 8 controls), failed a sex check (n = 0), were non-European population outliers (n = 4 cases, n = 7 controls) or were of southern European ancestry (n = 3 cases, n = 9 controls). For the UK/Dutch cohort, this left n = 410 cases and n = 767 controls.

Next, sample QC was performed on the entire UK/Dutch/French cohort (n = 2597). Samples with genotype missingness > 0.02 (n = 0) and those with heterozygosity rate 6 SD away from the mean (n = 6) were identified (Figure S2). Using LD pruned, variant QCed autosomal variants, relatedness was analysed using the KING relatedness tool[8]. Out of the 2597 samples, 18 unique samples were identified as being related through a 2nd degree or closer relationship (kinship coefficient > 0.0884). One of each pair was identified (n = 10 samples). Out of the 2597 samples, 1 duplicate was found; one of the pair was identified (n = 1). The 1000 Genomes Project (1KG) dataset was processed and filtered to only the pruned, variant QCed variants in our dataset; after merging, principal components analysis (PCA) was performed. Only samples of non-Finnish European ancestry were retained (n = 9 population outliers identified; Figures S3-4). All samples that failed the above sample QC were removed, leaving 2579 samples (n = 752 cases, n = 1827 controls). Using common, autosomal (excluding high-LD regions), LD pruned variants, ancestral PCs were generated using PC-AiR[9] for each sample in the case-control sample set (Figure S5). These were used later in the regression models.

#### Variant annotation and regions of interest

##### SpliceAI prioritisation criteria

Rare variants identified in BrS cases that potentially affected splicing of SCN5A transcripts, excluding likely pathogenic variants affecting the canonical splice donor and acceptor sites, were identified using SpliceAI predictions[10]. Candidate variants required a “double” SpliceAI prediction. For variants close to existing intron-exon junctions (exonic variants and intronic variants <50 bases from the junction), a minimum score of 0.2 for one of the predictions (donor loss, donor gain, acceptor loss, acceptor gain) was required. For deep intronic variants (>50 bases from intron-exon junctions), a prediction of both an acceptor gain and a donor gain was required (indicating the potential creation of a cryptic exon within the existing intron), with a minimum score of 0.2 for the first prediction and 0.1 for the second.

##### Epigenomic datasets and defining regions of interest

Epigenomic datasets were used to define regions of interest and help prioritise putatively deleterious genetic variants. All histone modification ChIP-seq data came from a left ventricle cardiac tissue sample from a female adult *Homo sapiens* in her 50s. This was provided by ENCODE[11–13] with codes as follows: ENCSR181ATL (human LV H3K4me3), ENCSR449FRQ (human LV H3K4me1) and ENCSR702OVJ (human LV H3K27ac). These datasets were generated by the Bradley Bernstein laboratory (Broad Institute). Two ATAC-seq datasets were used; one from a human cardiac left atrium sample (LA ATAC-seq)[14], the other from human pluripotent stem-cell derived ventricular-like cardiomyocytes (VLCM ATAC-seq)[15]. Promoter Hi-C from human induced pluripotent stem cell derived cardiomyocytes was used[16]. For cardiac transcription factor binding, the ReMap Atlas of Regulatory Regions ChIP-seq dataset was used (filtered to “cardiac”, “cardiac-muscle” and “cardiomyocyte” biotypes)[17]. For cardiac CTCF protein binding, the same dataset was used, except with the addition of a CTCF filter. In addition, a human cardiac myocyte CTCF ChIP-seq dataset from ENCODE was used[18].

Conserved, cardiac-accessible chromatin elements, E1-E25, of approximately 500 bp in length, were manually defined across the *SCN5A* TAD region (chr3:38.4-39.2 Mb). If the region was evolutionarily conserved (100 Vert. Cons. PhyloP), and had either a LA ATAC-seq or VLCM ATAC-seq peak, then it was defined as an element. This definition aims to broadly capture all possible relevant candidate regulatory elements for testing. For each element, ChIP-seq signals were considered; elements with only a CTCF ChIP-seq peak were excluded. A list of the intervals is provided in Table S2. Separately, CTCF-binding regions, C1-C18, were manually defined across the same *SCN5A* region. These regions were required to have all of: a human cardiac myocyte CTCF ChIP-seq peak (ENCODE), a cardiac tissue CTCF ChIP-seq peak (ReMap) and a JASPAR 2024[19] CTCF motif. A list of these intervals is provided in Table S3.

#### Transcription factor motif disruption prediction

Two different in-silico tools were used on candidate *cis*-regulatory element variants (SNVs and indels) to predict their impact on transcription factor binding motifs: motifbreakR[20] and homer2[21]. For motifbreakR, the p-value threshold was 1e-4, the IC method was used, and all motifs in MotifDb that had organism as “Hsapiens” were tested. Uniform background base probabilities were used (A=0.25, C=0.25, G=0.25, T=0.25). For homer2, wild-type and variant FASTAs were used as inputs, with a 10 bp flanking sequence either side of the variant base (all input FASTAs were 21 bp). For indels, bases were added/removed from the 5’/3’ ends to preserve the 21bp length. The HOMER/JASPAR database of 1,847 motifs was used.

#### ChromBPNet prediction

ChromBPNet models were trained using ventricle-like cells ATAC-seq data from the GEO dataset GSE146044. A bias model was first generated using a bias score threshold of 0.9, followed by training of the ChromBPNet models on the corresponding accessibility peaks and signals. The control scores for both the bias and ChromBPNet models passed the recommended quality thresholds, indicating satisfactory model performance.

### Genomic analyses

#### *Cis*-regulatory element and CTCF-binding site aggregate testing

Analyses were performed using the REGENIE (v3.3) tool[22]. First, a whole genome regression model was fitted using 528,851 genome-wide variants in REGENIE step one; only variants with a MAC > 100 were used. The first 10 ancestral PCs, sex (F-score derived), national_centre (UK/NL/FR) and SCN5A_positive were included as covariates in the regression models. An individual was "*SCN5A* positive" ({0,1}) if they possessed a nonsynonymous exonic or essential splice site *SCN5A* variant with a gnomAD FAF95_popmax < 1e-4 (gnomAD exomes ENST00000333535). By this definition, 19% of the cases were "*SCN5A* positive". The LOCO predictions generated during step one, were used as covariates in all downstream aggregate and single-variant testing analyses (REGENIE step two). This accounted for relatedness and population structure.

Aggregate variant testing (REGENIE step two) was performed on 25 conserved, cardiac-accessible chromatin elements, E1-E25, described earlier in the Methods. The covariates were as in step one, except SCN5A_positive was excluded. Only burden aggregate variant testing was performed, and only on singleton genetic variants (defined as MAC = 1; as opposed to "present", regardless of zygosity, in only one sample). The burden test referred to here and in all other analyses used the sum of alternative alleles method. A minimum cMAC was imposed for each mask (cMAC ≥ 5); 9 regions were ignored due to a low cMAC (cMAC < 5). A Bonferroni multiple-testing correction (shown as a red dashed line) was used: $\alpha_b = 0.05/16$, for the 16 tested regions. The nominal uncorrected significance level ($\alpha = 0.05$) was shown as a blue dashed line.

Aggregate variant testing was performed on 18 CTCF-binding regions, C1-C18; the intervals were defined in the "Epigenomic datasets and defining regions of interest" section. In addition, all of these regions were combined into one aggregate and tested. The covariates were as in REGENIE step one, except SCN5A_positive was excluded. The following aggregate variant tests were used: burden (assumes variants have the same direction and size of effect), SKAT (allows for variants to have different directions of effect), SKAT-O (omnibus test of burden and SKAT that tries to find the optimal combination of the two), ACAT-V (allows for variants to have different directions of effect and is powered even when only a small number of variants in the set are associated), ACAT-O (omnibus test of burden, SKAT, and ACAT-V that tries to find the optimal combination of the three). Only variants with a gnomAD-NFE allele frequency below 1% were included in the aggregate variant tests. For variance-component tests, variants with a MAC ≤ 3 were collapsed into a burden mask which was then included in the test instead of the individual variants. A minimum cMAC of 5 was imposed for each mask; 13 tested regions passed this with a cMAC ≥ 5. A Bonferroni multiple-testing correction (red dashed line) was used: $\alpha_b = 0.05/13$, for the 13 tested regions. The nominal uncorrected significance level ($\alpha = 0.05$) was shown as a blue dashed line.

#### Sliding window aggregate testing

Using REGENIE (step two), aggregate testing of singleton genetic variants across the *SCN5A* TAD (chr3:38488336-39139033; GRCh37) was performed using overlapping sliding windows. The burden test was used. An overlap of 20% was used; for the *i*-th window: $start_{i+1} = end_i - (20\% \times L_{window})$. The sliding window length was chosen to be 200 bp, as this approximates the length scale of an enhancer. Only singleton variants (as defined previously) with a CADD > 10 were considered. To be tested, the window had to have a cMAC ≥ 5. Covariates were as in step one, except SCN5A_positive was excluded. Out of 4066 windows, there were 22 windows that met the criteria to be tested.

#### Single-variant analysis

A single-variant analysis was performed on the *SCN5A* region (38.0-39.5 Mb) during REGENIE step two. This used a Firth-fallback logistic regression model. Only variants with a minimum MAC = 1 were considered. Covariates were as in REGENIE step one, including SCN5A_positive. The output from step two was filtered to only variants with an $AAF_{gnomAD\text{-}popmax} < 1\%$. A Bonferroni multiple-testing correction was used.

#### Monte-Carlo overlap enrichment experiments

In order to determine whether BrS case singleton genetic variants (both SNVs and small indels; singletons defined as $MAC_{cases} = 1$ and $MAC_{controls} = 0$) intersect the cardiac-specific accessible chromatin elements in the *SCN5A* region more often than would be expected by chance, a Monte-Carlo permutation testing approach was utilized, as follows:

1. The BrS case singleton variants were positioned randomly in the region (sampling from a uniform probability distribution across the region).
2. The number of intersections between these SNVs/indels and the cardiac-specific accessible chromatin elements was recorded.
3. Steps (1) and (2) were repeated 1000 times ($n_{permutations}$).
4. A histogram of the number of intersections from each trial was generated—this acted as the empirical null distribution, constructed by simulation.
5. The number of intersections observed when the BrS case singletons were in their actual observed genomic positions was computed and this value was compared with the expected value from the empirical null distribution.

This was implemented using BEDTools[23]. The permutation testing *p*-value was computed as the proportion of sampled permutations whose value was more extreme than the observed number of intersections; if the observed number of intersections was greater than all simulated number of intersections then the *p*-value was taken to be $p < 1/n_{permutations}$. The estimated effect size was computed as $o/e$, where $o$ is the observed number of intersections and $e$ is the expected number of intersections (computed as the mean of the empirical null distribution). The same experiment was also performed using BrS-associated common variants. Concretely, the set of 8 conditionally independent genome-wide significant SNPs, identified via the Barc *et al.* BrS GWAS[24], as well as all SNVs in LD ($r^2 > 0.6$) with them, was used.

#### Structural variant calling and mobile element insertion conditional analysis

The Delly2 and Manta SV callers were applied to the $n$ = 2515 UK, NL and FR BAMs (cases and controls; post-sample QC); the IndexCov SV caller was applied to the corresponding BAIs (BAM index files). The region considered was chr3:38.0-39.5 Mb (GRCh37). Delly2 called $n$ = 43 structural variants in the region. Manta SV calls were collapsed using Truvari, in order to properly call common/low-frequency SVs. After collapsing, Manta called $n$ = 695 SVs. Manta called a common SCN5A mobile element insertion (MEI). Targeted SV calling for this MEI was performed on the same set of BAMs using an SV caller specifically designed for the detection of mobile elements—Mobile Element Locator Tool (MELT)[25].

A conditional analysis was performed between the MEI and the 8 conditionally independent Barc et al. (2022) BrS GWAS SNPs. A single-variant analysis across chr3:38.0-39.5 Mb was performed using REGENIE step two, as previously described, except with no AAF filter. This was repeated, this time with the addition of the MEI to the conditioning list. The change in significance of the 8 conditionally-independent BrS GWAS SNPs was considered.

#### Differential singleton kernel density estimation

Kernel density estimation was used to compute the difference between the spatial (genomic position) density of case ($MAC_{cases}$ = 1, $MAC_{controls}$ = 0) and control ($MAC_{cases}$ = 0, $MAC_{controls}$ = 1) singleton genetic variants across the *SCN5A* gene and directly upstream and downstream (chr3:38548315-38748094; GRCh37). This was implemented using the density function in R. A triangular smoothing kernel function was used; the smoothing bandwidth was 0.20 times the default.

#### Gene-centric aggregate testing

Gene-centric aggregate variant testing was performed using different masks. Using ANNOVAR's variant annotation: 1) a UTR mask (UTR = {5' UTR, 3' UTR}) was generated for all coding and noncoding genes in the *SCN5A* region; 2) an exonic ncRNA mask was generated for all of the noncoding RNA genes in the *SCN5A* region; 3) a nonsynonymous coding variant mask was generated for all protein-coding genes in the *SCN5A* region, where nonsynonymous = {essential splice site variant, frameshifting indel, start loss, stop gain, non-frameshifting indel, nonsynonymous SNV}. Using VEP, variants in the region were annotated with spliceAI scores; a splice mask containing all variants with a spliceAI score > 0.1 (any of AG, AL, DG or DL) was generated for all genes in the *SCN5A* region.

The covariates were as in REGENIE step one, except SCN5A_positive was excluded. The following aggregate variant tests were used: burden, singleton burden, SKAT, SKAT-O, ACAT-V, ACAT-O. Only variants with a gnomAD-NFE allele frequency below 1% were included in the aggregate variant tests (other than the singleton burden). For variance-component tests, variants with a MAC ≤ 3 were collapsed into a burden mask which was then included in the test instead of the individual variants. A minimum cMAC of 5 was imposed for each mask in order to be tested. A Bonferroni multiple-testing correction (red dashed line) was used: $\alpha_b = 0.05/n$, where $n$ is the number of genes tested; this differed between masks. Specifically for the nonsynonymous mask: genome-wide significant independent BrS GWAS SNPs were conditioned upon (Barc et al.[24] and Makarawate et al.[26]); the *SCN10A*-short isoform was included in the analysis by performing aggregate coding variant testing on the last 7 exons of *SCN10A* (NM_006514, exons 21-27) and only variants with a gnomAD-NFE allele frequency below 0.1% were included. These results are shown in Figure S7. Separately, we also burden tested nonsynonymous variants in *SCN10A* and *SCN10A*-short specifically again, this time using a lower AAF cut-off: $AAF_{gnomAD\text{-}NFE} < 1\times10^{-4}$. These results are mentioned in the main text.

#### Replication of associations: Health in Code genetic analysis

To replicate BrS associations in non-coding regions of SCN5A, we utilised an independent BrS referral cohort from the genetic testing company Health in Code S.L., Spain. The cohort included 483 probands that were referred for BrS genetic testing and were sequenced on a large gene panel that included coverage of all *SCN5A* introns, enabling replication of associations at internal non-coding regions of *SCN5A* (E14 CRE and intron 1 low frequency variants). As the gene panel assay does not cover introns in *SCN10A* or regions up and downstream of *SCN5A*, other associations could not be tested for replication (e.g. E22, E23 CREs).

For controls, we utilised 12,259 probands with non-channelopathy conditions that were sequenced on the same panel and are expected to represent an equivalent cohort with respect to ancestral background. These individuals were referred for genetic testing for cardiomyopathies, aortic diseases, and dyslipidaemias. Data from gnomADv4 was used as secondary control cohorts (global and non-Finnish European) but these datasets are expected to be less well matched to the BrS cases with respect to ancestry, as the Health in Code referral cohorts are enriched for individuals from Southern Europe[27].

The phenotypes of these cases were determined by the respective referring centres. As this is a referral cohort, it is expected to include a lower proportion of definitively diagnosed BrS cases, as evidenced by the relatively low prevalence of pathogenic/likely pathogenic variants (9.04%), compared to a typical European ancestry BrS cohort of ~20%. As a result, case-control association signals would be expected to be somewhat diluted compared to our primary BrS cohort.

The NGS customized sequencing library was enriched using a specificity hybridization probe kit SureSelect XT HS Low input (Agilent) and the obtained fragments were sequenced in paired-end mode (2x150bp) on the NovaSeq X Plus Sequencing System (Illumina). Sequencing data analysis was performed using a proprietary bioinformatics pipeline comprising sample demultiplexing, alignment refinement and adjustment, variant calling, annotation, and sequence quality control. The read depth (number of times that a base was sequenced by independent reads) of every nucleotide of genes related to the referring phenotype was >30× (mean 250× to 400×).

#### Effect on ECG endophenotypes in the UK Biobank

The UK Biobank (UKB) is a large-scale resource that contains extensive data on biological characteristics, health, and lifestyle. Between 2006 and 2010, around 500,000 individuals aged 40–69 were recruited across the United Kingdom. At enrolment, participants completed standardized clinical assessments, provided detailed information through health and lifestyle questionnaires, and contributed biological samples for laboratory testing and genetic analyses. Follow-up is conducted through repeat assessments in subsets of participants, as well as through linkage to national electronic health records (EHR).

Whole-genome sequencing (WGS) data were internally analysed using the DRAGEN pipeline, and the processed outputs were made available to researchers in PLINK2 format. Detailed descriptions of the general quality control (QC) procedures applied at both the variant and sample levels have been published previously[28]. Ethical approval for the UK Biobank resource was granted by the UK Biobank Research Ethics Committee, and all participants provided informed consent. The analyses presented here were performed under the approved UK Biobank application number 176602.

Two types of ECG data are available in the UK Biobank as raw XML files: resting ECGs and exercise ECGs. The exercise ECG recordings were obtained between 2006 and 2013 using a 4-lead system (CAM-USB 6.5, Cardiosoft version 6.51). Each recording lasted 7 minutes and 15 seconds, with the initial 15 seconds representing a pre-test “resting” phase, which is used in this study. Resting ECGs were collected between 2014 and 2024 using a standard 12-lead system (GE Cardiosoft version 6), with each recording spanning 10 seconds.

The following ECG parameters were extracted from the raw XML files: heart rate (HR), PR interval, RR interval, QRS duration, and QTc interval. The QC procedures for “minimal” and “strict” filtering are summarized in Table ST7. In this analysis, the “minimal” QC pipeline was applied, where ECG parameters were adjusted for the use of beta-blockers and calcium antagonist medication. The 12-lead and 4-lead ECG recordings were combined, tagging the source of ECG to be used later as a covariate. When multiple ECG measurements were available for the same individual, preference was given to 12-lead ECG recording. If multiple measurements existed within the same source, priority was given to those meeting strict sample and ECG QC criteria. If still multiple remained, the earliest recorded ECG was selected. The minimal QC procedures resulted in 125,981 HR measurements, 123,270 QTc measurements, 126,153 PR measurements, 125,033 QRS measurements, and 124,512 RR measurements for individuals in the UK Biobank.

Genome-wide association studies (GWAS) were performed using REGENIE (v. 4.1.2)[22]. In step 1, a whole-genome ridge regression model was fitted for each ECG phenotype using a pruned subset of the genotype data (555,693 variants), generating genomic predictions. Both the raw and inverse rank-normal transformed (IRNT) values of each ECG parameter were analysed. In addition, analyses were performed both in the full cohort and in a subset restricted to individuals of European ancestry. The predictions were subsequently used in step 2 for quantitative single variant association testing with WGS data. All analyses were adjusted for Age, $Age^2$, Sex, Batch, UKB assessment centre region, ECG source, and the first 20 principal components. Variants with a minor allele count (MAC) < 10 were excluded from the analyses.

### Cis-regulatory element functional validation

#### Generation and preparation of hiPSC-CMs for transfection experiments

The hiPSC line LUMC0099iCTRL#04 (generated by the iPSC core facility of Leiden University Medical Center and registered at LUMCi004-A · Cell Line · hPSCreg) was maintained in undifferentiated state in presence of mTeRS1 medium on Matrigel Matrix-coated plates, and differentiation started when it reached 85-95% confluence. Cardiomyocyte differentiation was performed as described by Maas et al.[29] with the following modifications. Differentiation started in RPMI 1640 medium containing 2% B27 supplement without insulin (both by Gibco/Thermo Fisher Scientific) with the addition 213ug/ml of Lascorbic acid 2-phospahte (Sigma-Aldrich/Merck) (hereafter indicated as RPMI/B27−) and 6 µM CHIR99021 (Selleck Chemicals) for 3 days followed by 3-days treatment with 2 µM Wnt-C59 (MedChemExpress; Wnt signaling pathway inhibitor) in RPMI/B27− medium. On day 8 of differentiation the medium was switched to RPMI 1640 containing 2% B27 supplement with insulin and without L-ascorbic acid 2-phosphate (hereafter indicated as RPMI 1640/B27+) until day 30, with medium change every 3-4 days. A 7-8 day metabolic-selection that enriches the culture for cardiomyocytes was then started by switching the medium to RPMI 1640 without glucose (Gibco/ThermoFisher Scientific) supplemented with 500 µg/mL bovine serum albumin (SigmaAldrich/Merck) and 8 mM Na-L-lactate (Sigma-Aldrich/Merck) with medium change every other day (adapted from Tohyama S. et al.[30]). The culture was then switched to RPMI 1640/B27+ for 1 day before proceeding to dissociation and reseeding for transfection experiments. Dissociation of hiPSCcardiomyocytes culture was performed by incubation in presence of StemPro Accutase (Gibco/ThermoFisher Scientific) for 45minutes at 37ºC and 5%CO2, followed by gentle dissociation by pipetting, dilution of the dissociation reagent with RPMI 1640/B27+ medium before centrifugation and resuspension of the hiPSC-cardiomyocytes pellet in RPMI 1640/B27+ medium. Cardiomyocytes were seeded at 10.5x10^4 cells/cm2 in RPMI 1640/B27+ medium and transfected 7 days later.

#### Cell culture and transfection luciferase assays

The RE5 (which contains E14), E17, E22, and E23 fragments were cloned into a modified pGL2-Basic plasmid containing an SV40 minimal promoter and an adjusted multiple cloning site for *in vitro* analysis by a transfection luciferase assay. hiPSC-CM cultures were maintained as described above. Standard transfections (day 0) of hiPSC-CMs with 200 ng of reporter construct in 48 well-plates, were carried out using ViaFect transfection reagent (Promega E4981), using 1:6 reagent:DNA ratio. On day 1, cells were washed in RPMI 1640/B27+ medium 24 hours after transfection. On day 4, cells were lysed using Renilla luciferase assay lysis buffer (Promega, E291A-C) and luciferase activity was measured. Luciferase measurements were performed using a GloMax Explorer (Promega, GM3500). During the measurement, 100 µL D-Luciferin (p.j.k, 102111) was injected (150 µL/second) followed by a 1 second delay and 5 seconds of measurement. Transfections were carried out at last four times and measured in duplicates. Statistics were performed using one-way ANOVA, followed by Dunnett's multiple comparisons test for RE5, Welch's ANOVA followed by Dunnett's multiple comparisons test for E17 and E22, and Kruskal-Wallis followed by Dunn's multiple comparisons test for E23.

#### Fragment coordinates (hg38)

The fragment coordinates are as follows:

- RE5: chr3:38579568-38580860
- E17: chr3:38633969-38634738
- E22: chr3:38723560-38724959
- E23: chr3:38725399-38726531

### Structural variant functional validation

#### Patient recruitment and deletion identification

The patient was recruited following a medical consultation and presented with a type I Brugada pattern on their ECG after an Ajmaline test. DNA was extracted from the patient's peripheral blood, and 1.1 µg was used to prepare a library for whole-genome sequencing (WGS) along with 349 other patients affected by the same syndrome, using Illumina's TruSeq DNA PCR-Free Library Preparation Kit, according to the manufacturer's instructions. After normalization and quality control, the qualified libraries were sequenced on an Illumina HiSeqX5 system (Illumina Inc., CA, USA) with 150 bp paired-end reads. Each sample achieved a minimum sequencing depth of 80% and ≥20x coverage. The WGS pipeline followed the Broad Institute GATK "Best Practices" guidelines. Briefly, reads were mapped to the human genome GRCh37 using bwa-mem, duplicates were marked using Picard, and GATK was used for realignment and recalibration of reads (broadinstitute.org/gatk/guide/bestpractices.php). Structural variants were called using the LUMPY framework, with structural variants detected through lumpy-sv express v0.2.13. Depth profiles were plotted using lowresbam2raster from the jvarkit package (http://dx.doi.org/10.6084/m9.figshare.1425030).

#### Generation of hiPSCs and ventricular-like differentiation

Human induced pluripotent stem cells (hiPSCs) were derived from peripheral blood mononuclear cells and reprogrammed using the Sendai virus within the iPSC platform at the University of Nantes. The hiPSCs were obtained from a man in his 50s, previously described[31], who exhibited no clinical symptoms and had a normal ECG. The hiPSCs were maintained on stem cell-specific Matrigel-coated plates (0.1 mg/mL BD Bioscience) and cultured in StemMACS iPS-brew XF medium (Miltenyi Biotec). The hiPSCs were passaged every 3 to 4 days using the Gentle Cell Dissociation Reagent (StemCell Technologies). To generate ventricular cardiomyocytes (CMs), hiPSCs were grown as a monolayer in StemMACS iPS-brew medium supplemented with the Y-27632 ROCK inhibitor 1/1000 (Stemcell Technologies). When hiPSCs reached 70-80% confluency, they were differentiated into hiPSC-CMs using the previously described 'Matrix sandwich' method[32]. From day 10 to day 13 of differentiation, the beating hiPSC-CMs were purified using a glucose deprivation protocol with glucose-free RPMI1640 medium (Life Technologies)[33]. After this initial depletion, cells were detached and re-seeded in a new plate, and a second depletion was carried out from day 14 to day 17. On day 17, hiPSC-CMs were cultured in RPMI1640 medium supplemented with complete B27 (Life Technologies), 1X L-glutamine, and 1% NEAA, with medium changes every two days until the end of differentiation on day 30.

#### Generation of the isogenic model containing the 10 kb deletion on hiPSCs

The 10 kb deletion in the control hiPSC cell line was performed using the previously described Alt-RTM CRISPR-Cas9 system (Caillaud et al., 2022[34], Appendix 2). Briefly, guide RNAs (gRNAs; Integrated DNA Technologies (IDT)) were designed to target the deletion ends (sequence: gRNA1: ccaccatctaagatgcctcc and gRNA2: tatagtatgtgccttaactc). hiPSCs were transfected with the Alt-R CRISPR-Cas9 complex using the Amaxa P3 Primary Cell 4D-Nucleofector® X Kit (Lonza, #V4XP-2024). ATTO488-positive cells were sorted into 96-well plates to obtain clonal populations using a FACSMelody™ (BD Biosciences). This initial genome editing generated two clones, referred to hereafter as hiPSC-CM-V-del Clone 1 +/- and hiPSC-CM-V-del Clone 2 +/-. Multiple rounds of genome editing on these heterozygous clones were subsequently attempted but did not yield homozygous clones for the deletion (0/402). For genotyping the isogenic models, hiPSCs were lysed using 30 µL of QuickExtract solution (Epicentre), and DNA was extracted following the manufacturer's instructions. PCRs were conducted to genotype the 10 kb region (Primers: F: atctccttcagtgggtgtgc / R:

tgtgtttgcgtagtttccaaag / intraF: cgacgttttcaaagtgctga) followed by Sanger sequencing (Eurofins Genomics).

#### Transcriptomic analyses of the isogenic hiPSC-CM-V model

Total RNA from the hiPSC-CM-V-10Kb-/+ clone 1 and hiPSC-CM-V-10Kb-/+ clone 2, derived from 7 and 5 differentiations respectively, and total RNA from hiPSC-CM-V WT derived from 10 differentiations, was isolated using the NucleoSpin RNA Kit (Macherey-Nagel) following the manufacturer's instructions. The quality and integrity of RNA samples were assessed using the 2100 Bioanalyzer and the RNA 6000 Nano LabChip II Kit (5067-1511, Agilent Technologies). RNA libraries were prepared by the GenoBiRD platform according to their published method[35] and sequenced on the NovaSeq 6000 (Illumina). Primary and secondary analysis, including demultiplexing, alignment to the GRCh38 reference genome, quality control, counting steps, principal component analysis, and differential expression between the two conditions, were performed using the Snakemake pipeline developed by the GenoBiRD platform[35].

#### Electrophysiological studies

Four differentiations of hiPSC-CM-V-WT, four differentiations of hiPSC-CM-V-del Clone 1 +/-, and three differentiations of hiPSC-CM-V-del Clone 2 +/- at days 32–35 of differentiation were dissociated into single cells using Accutase (Sigma) and resuspended after washing (300,000 cells/mL) in a solution composed of 50% RPMI 1640 + complete B27 and 50% extracellular buffer containing in mM: 140 NaCl, 4 KCl, 2 $CaCl_2$, 1 $MgCl_2$, 5 glucose, and 10 HEPES (pH 7.4, osmolarity 298 mOsm) supplemented with 5 µM nifedipine. The cell currents were recorded using the high-throughput patch-clamp system by Nanion (SyncroPatch 384 PE, Nanion, Munich, Germany), which allows for recording the currents of 384 cells in parallel. Single-hole, low-resistance chips (3–5 MΩ) were used. The intracellular solution contained in mM: 10 CsCl, 110 CsF, 10 NaCl, 10 Ethylene Glycol Tetraacetic Acid (EGTA) and 10 HEPES (pH 7.2, osmolarity 280 mOsm). After cell catching, sealing, whole-cell formation, liquid application, recording, and data acquisition were all performed sequentially and automatically.

Whole-cell experiments were performed at a holding potential of −100 mV at room temperature (18-22°C). Currents were sampled at 20 kHz. Current density was measured by a depolarization to +10 mV for 50 ms. repeated 10 times with a 5-second interval to ensure current stability. Activation curves were built by 2000 ms-lasting depolarisations from -150 mV to 45 mV using 5 mV increment steps. Steady-state inactivation curves were conducted by using a 50 ms-lasting depolarization step to -20 mV after the activation steps. Pulses were applied every 10 sec, time during which cells were mostly hold at -100 mV before prepulse and pulse application. Both activation and inactivation curves were fitted using Boltzman equations to evaluate half-potentials of activation and inactivation, respectively.

The data were analyzed in R using internally developed scripts, with representations and statistical tests (Mann-Whitney t-test) generated in Graphpad 8 software.

### Splice variant functional validation

#### Minigene assays

We studied 1 variant using a minigene method previously reported by one of our labs[36]. Briefly, the respective exon and 100 nucleotides of flanking intronic sequence were amplified from genomic DNA (see Table S4 for primers). The amplicon and minigene vector pET01 (MoBiTec GmbH) were digested with NotI and SalI (New England Biolabs), the vector further digested with Calf Intestinal Phosphatase (Promega), and then gel extracted and purified (Qiagen). The insert and vector were ligated with T4 ligase (New England Biolabs), transformed into DH5 cells, and plated on ampicillin agar plates. Colonies were picked and grown overnight with 1 mg/mL ampicillin. DNA was isolated using the Miniprep kit (Qiagen) and Sanger sequenced to yield a wild-type minigene vector. The variant was introduced with the QuikChange Lighting Multi kit (Agilent) per manufacturer's protocol to produce the variant plasmid. Human Embryonic Kidney (HEK293) cells at 50% confluency were transfected with 500 ng of the wild-type and minigene plasmids using FuGENE6 (Promega) per manufacturer's protocol. After 48 hours, HEK293 cells were harvested and RNA isolated using the Qiagen RNeasy Minikit. A primer specific to pET01 was used for reverse transcription with the SuperScript III System (Invitrogen). PCR of cDNA using the GoTaq PCR Master Mix (Promega) produced amplicons that were fractionated on a gel, extracted, and Sanger sequenced to determine splicing consequence.

#### CRISPR-Cas9 iPSC experiments

We studied one deep intronic *SCN5A* variant using CRISPR-Cas9 gene editing of iPSCs. Guides were designed using the CRISPOR online tool[37] and cloned into SpCas9-2A-GFP[38] (Addgene #48138, a gift of Feng Zhang) as previously described[36]. The guide plasmid and a homology directed repair template bearing the variant of interest and PAM-site variant were electroporated with a Neon Transfection System (ThermoFisher MPK5000) into previously characterized control cells from a healthy donor[39]. GFP+ cells were sorted 48 hours post transfection with a BD Fortessa 5-laser instrument. Individual colonies were genotyped with variant-specific primers and heterozygous clones were chosen for further studies. iPSCs were grown on Matrigel (BD Biosciences) coated plates in mTeSR-Plus media (STEMCELL). Cardiac differentiation was completed with a monolayer chemical method as previously described[40]. To determine splicing outcomes, cells were treated with vehicle dimethylsulfoxide or cycloheximide (100 M) for 6 hours. Immediately after, RNA was isolated from iPSCs or cardiomyocytes using the RNeasy Minikit protocol (Qiagen). We performed RT-PCR as above with variant specific primers, after which amplicons were separated by gel electrophoresis, extracted, purified, and Sanger sequenced to determine splicing consequences.

### Supplemental Figures

#### Figure S1: Variant QC metrics

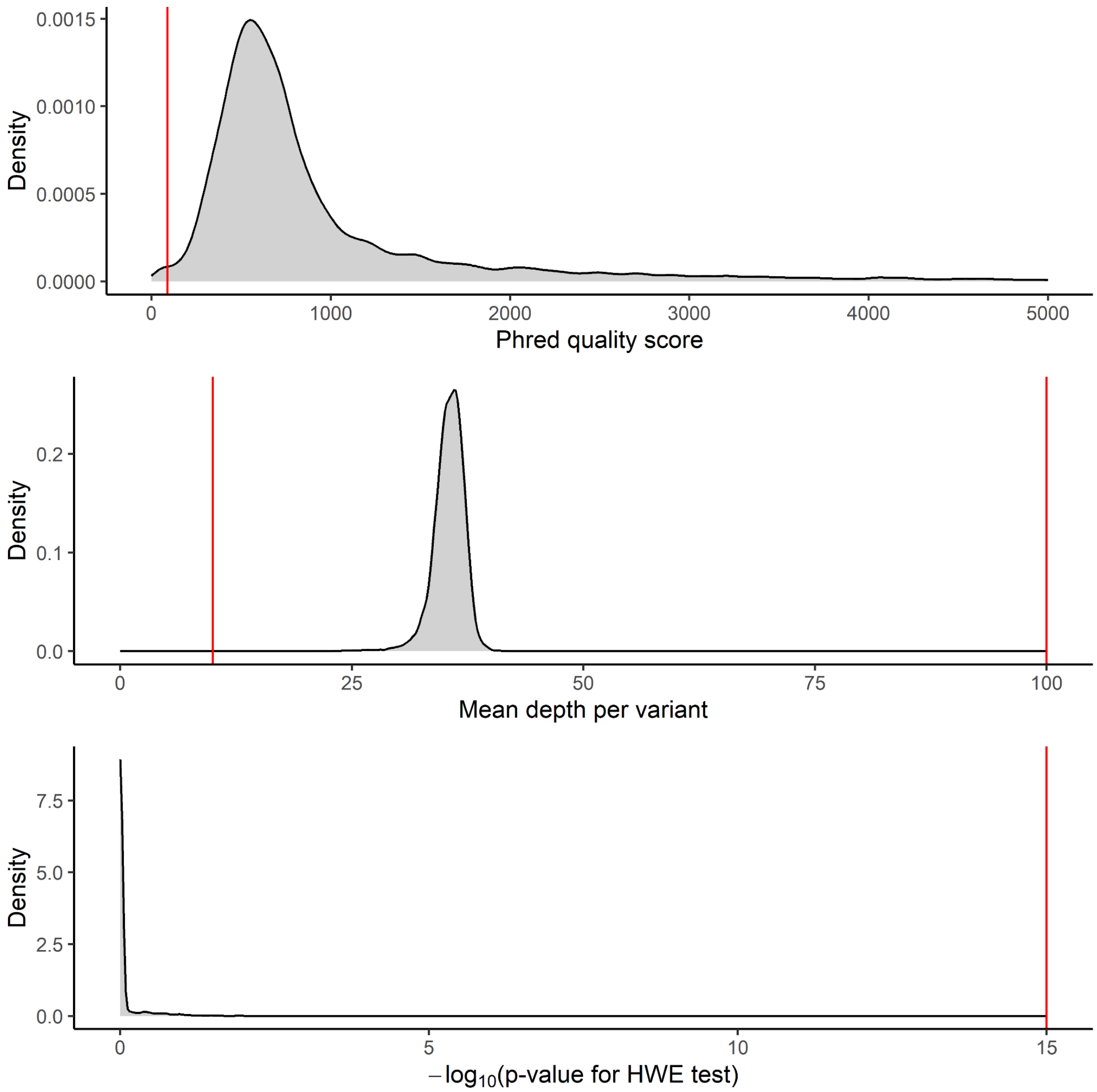


**Figure S1 | Variant-level QC metrics.** Histograms of variant Phred quality scores, mean depths and Hardy-Weinberg Equilibrium test p-values. Red vertical lines indicate the QC thresholds applied.

### Figure S2: Heterozygosity and missingness sample QC

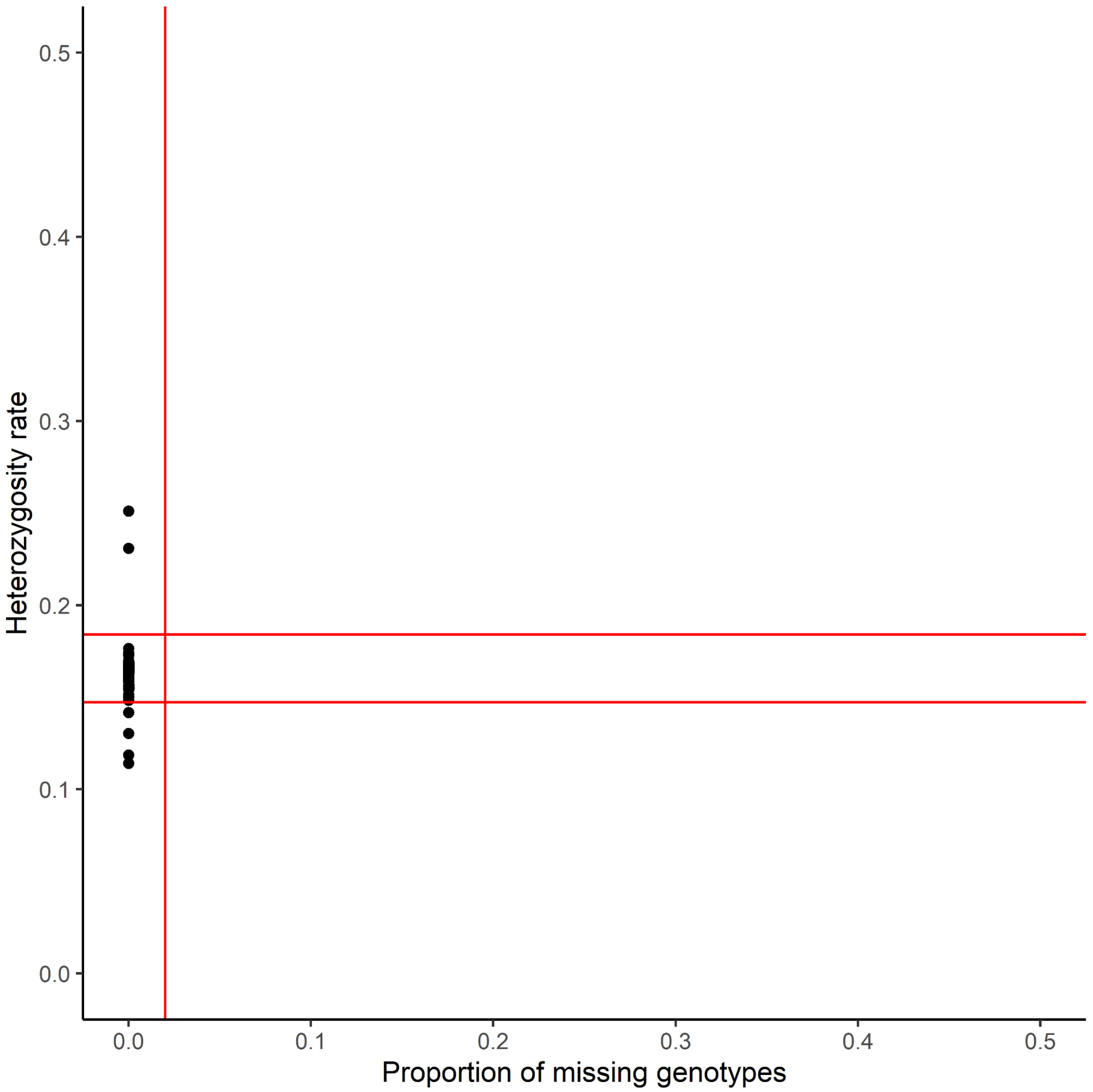


**Figure S2 | Heterozygosity and missingness sample QC.** Plot of sample-level heterozygosity rate versus genotype missingness. Red lines indicate the QC thresholds applied.

### Figure S3: Sample ancestry (global)

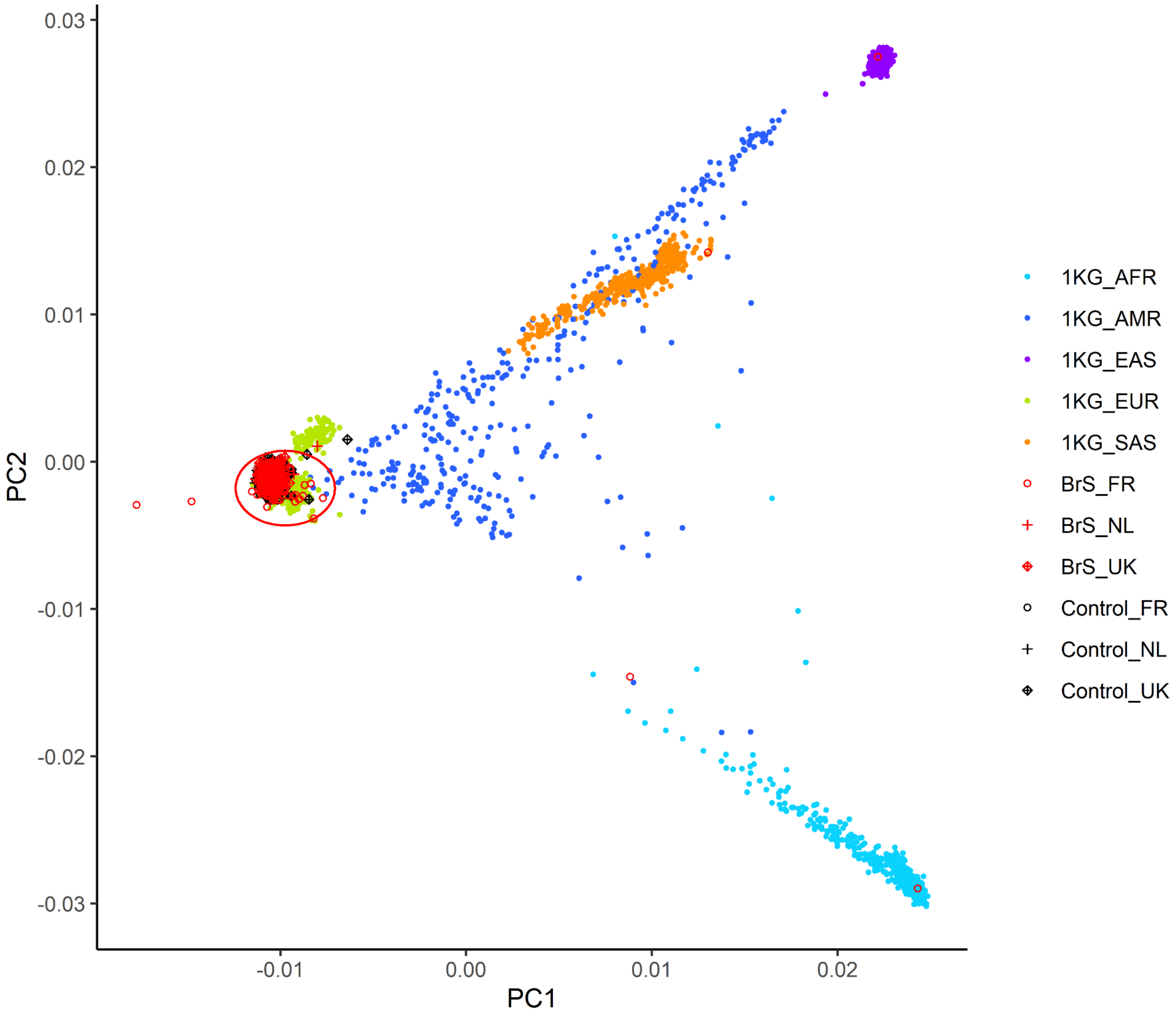


**Figure S3 | Sample ancestry (global).** PCA plot of case and control samples with 1000 Genomes Project samples as reference, illustrating ancestry of the cohort at the global level. Red ellipse indicates the samples taken forward.

### Figure S4: Sample ancestry (European)

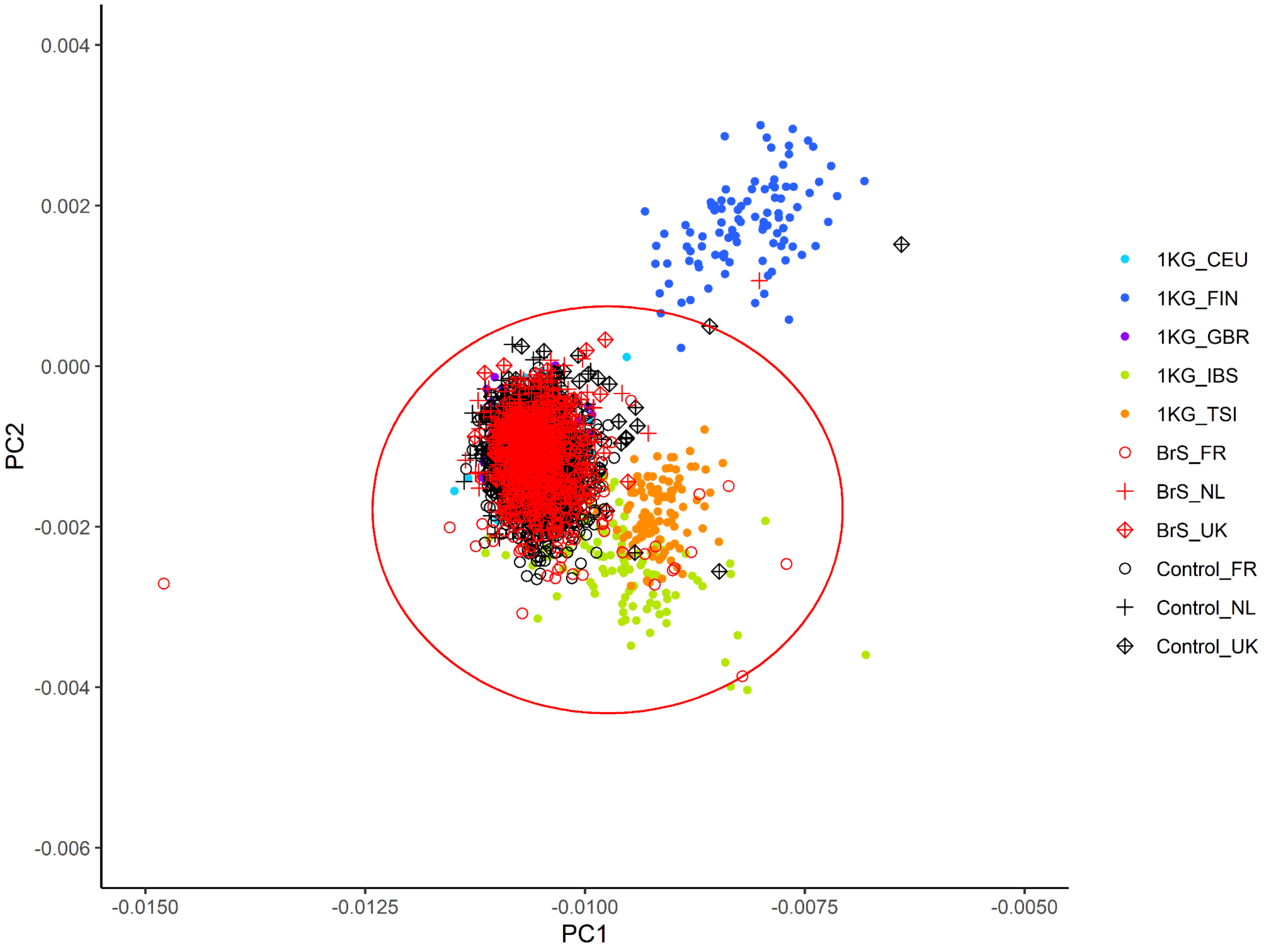


**Figure S4 | Sample ancestry (European).** PCA plot of case and control samples with 1000 Genomes Project samples as reference, illustrating ancestry of the cohort at the European level. Red ellipse indicates the samples taken forward.

### Figure S5: Case-control ancestral PCs

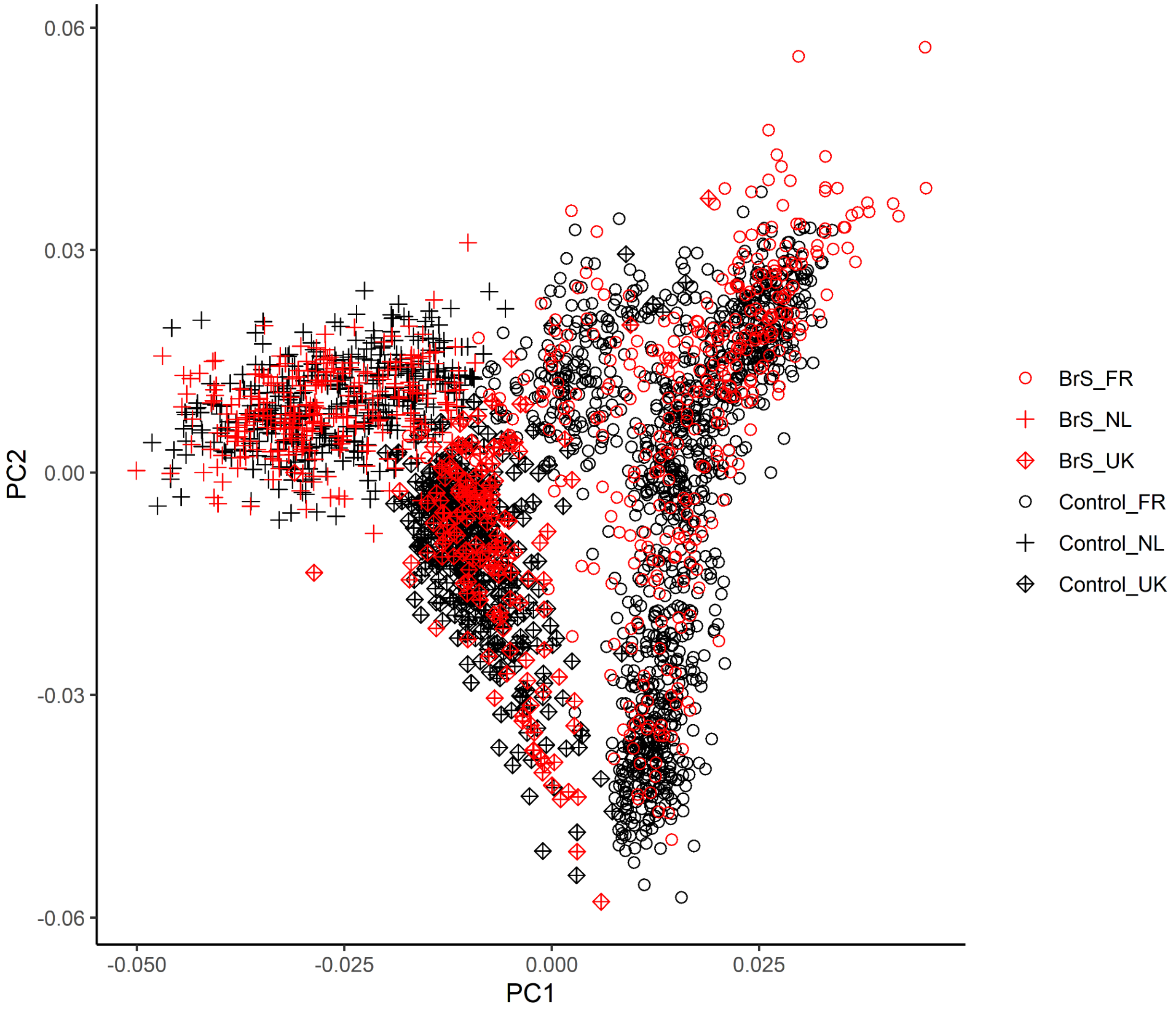


**Figure S5 | Case-control ancestral PCs.** PCA plot of case (red) and control (black) samples, showing good overlap (case-control ancestry match).

### Figure S6: Functional studies of potential splice-altering variants

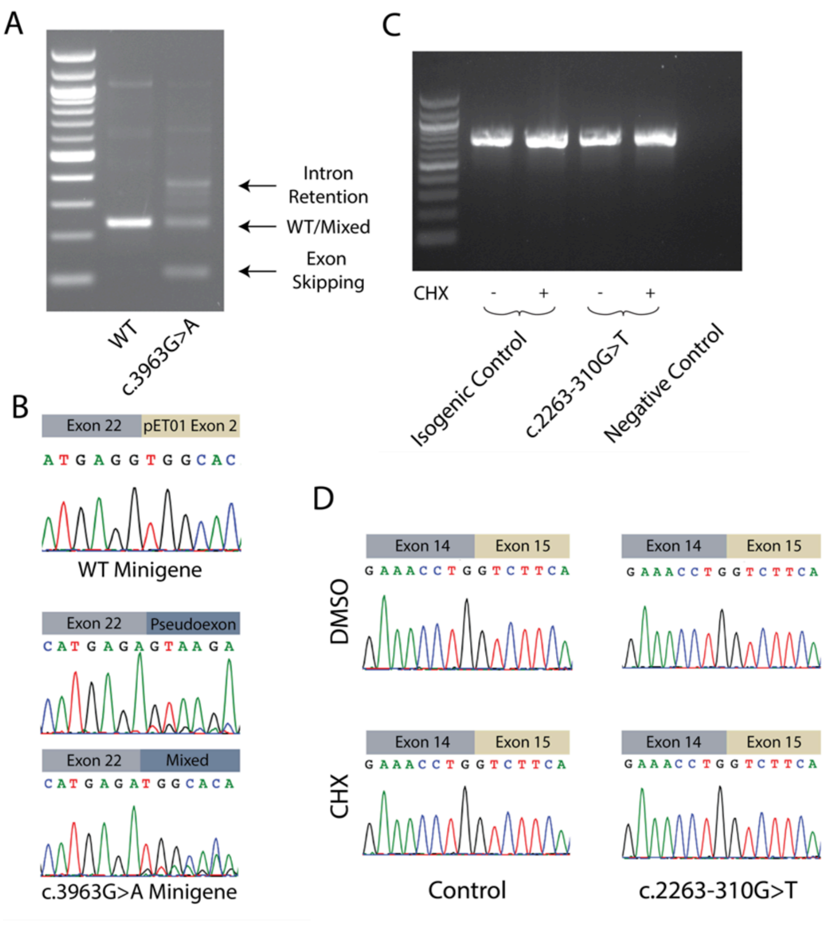


**Figure S6 | Functional studies of potential splice-altering variants.** A) Gel electrophoresis of Reverse Transcription Polymerase Chain Reaction (RT-PCR) amplicons of RNA isolated from HEK cells transfected with WT (c.3963G) and variant (c.3963A) minigene vectors. A single band is observed in the WT lane and multiple bands are observed in the variant lane. B) Sanger sequencing of gel-extracted bands from panel A revealed that canonical splicing occurs with the WT minigene, spanning exon 22 into minigene pET01 exon 2. Introduction of the variant introduces retention of a pseudoexon, a mixture of aberrant splicing and WT, and an exon-skipping event. C) Gel electrophoresis of RT-PCR amplicons of RNA isolated from iPSC-CMs CRISPR-edited to include c.2263-310G>T and an isogenic WT control. CHX indicates treatment with 100 M cycloheximide (CHX), an inhibitor of Nonsense-Mediated Decay. Negative control indicates no input to the PCR. D) Sanger traces of gel extracted products from panel C. Canonical splicing between exon 14 and exon 15 is observed for both WT and c.2263-310G>T cells, irrespective of CHX addition.

### Figure S7: CTCF, UTR and exonic ncRNA genetic variation

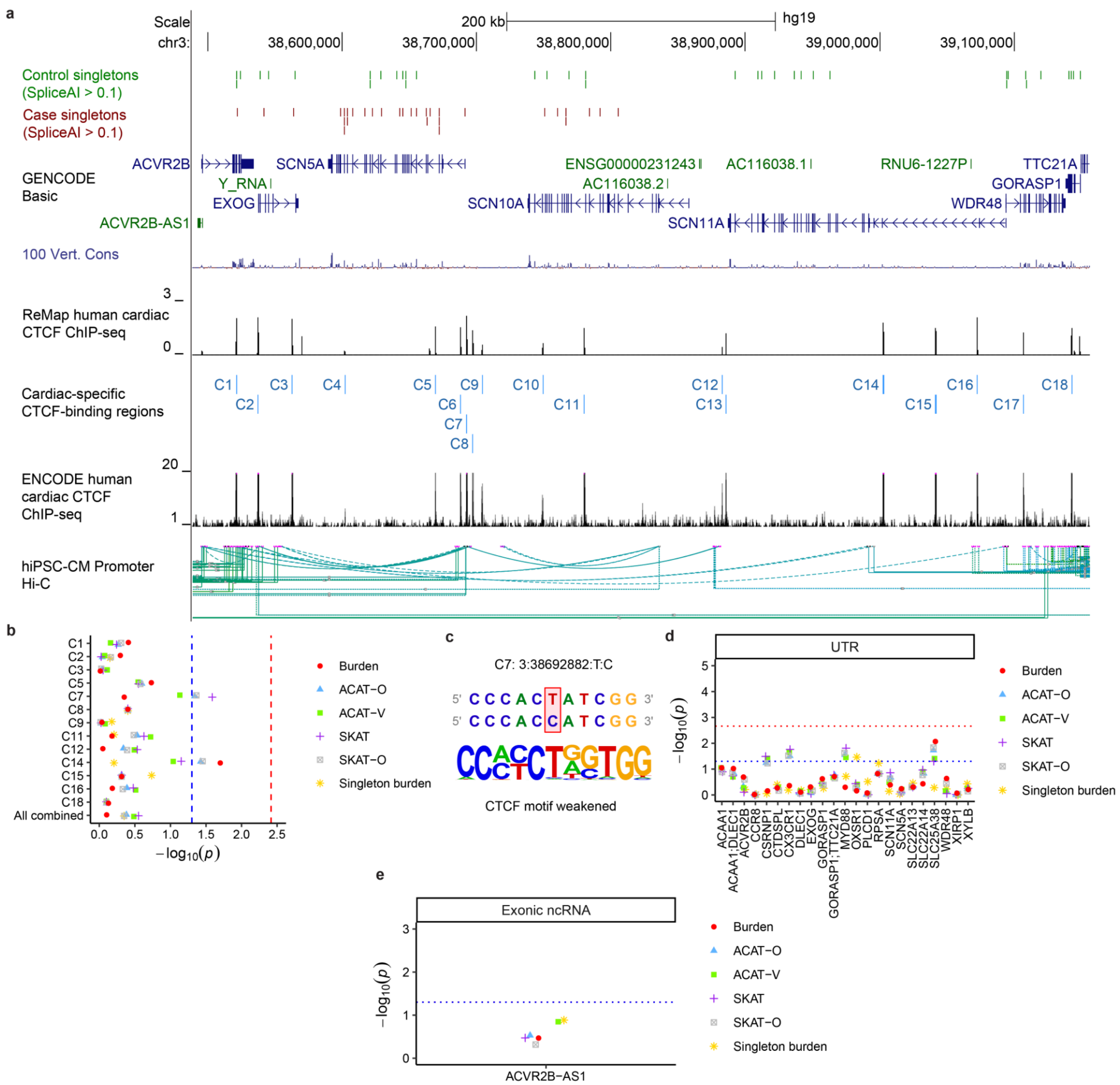


**Figure S7 | CTCF, UTR and exonic ncRNA genetic variation.** A) View of the SCN5A TAD with the protein-coding genes (blue) and ncRNA genes (green) shown. Case and control singletons that have a SpliceAI score > 0.1 are shown as red and green dashes respectively. Regions of CTCF protein binding are shown (ReMap human cardiac ChIP-seq and ENCODE human cardiac ChIP-seq) along with the manually defined cardiac-specific CTCF-binding regions

(C1-C18; light blue). Physical connections between promoters and points of chromatin are shown by the hiPSC-CM promoter Hi-C.

**Figure S7 |** *(Continued.)* B) Rare variant burden testing of the CTCF-binding regions ($AAF_{gnomAD\text{-}popmax}$ < 1%; blue dashed line: nominal significance, α = 0.05; red dashed line: Bonferroni-corrected threshold, $\alpha_b$ = 0.05/13). C) Case singleton variant predicted to alter a CTCF motif with flanking reference (top row) and alternative (bottom row) genomic sequences; affected base(s) are highlighted in red. The aligned sequence logo of the CTCF motif is shown underneath. D) Rare variant burden testing of the combined 3' and 5' UTRs of genes in the SCN5A TAD ($AAF_{gnomAD\text{-}popmax}$ < 1%; blue dashed line: nominal significance, α = 0.05; red dashed line: Bonferroni-corrected threshold, $\alpha_b$ = 0.05/23). E) Rare variant burden testing of the only ncRNA gene in the SCN5A TAD that met the criteria for testing ($AAF_{gnomAD\text{-}popmax}$ < 1%; cMAC ≥ 5 for inclusion; blue dashed line: nominal significance, α = 0.05).

### Figure S8: Read evidence for SV1

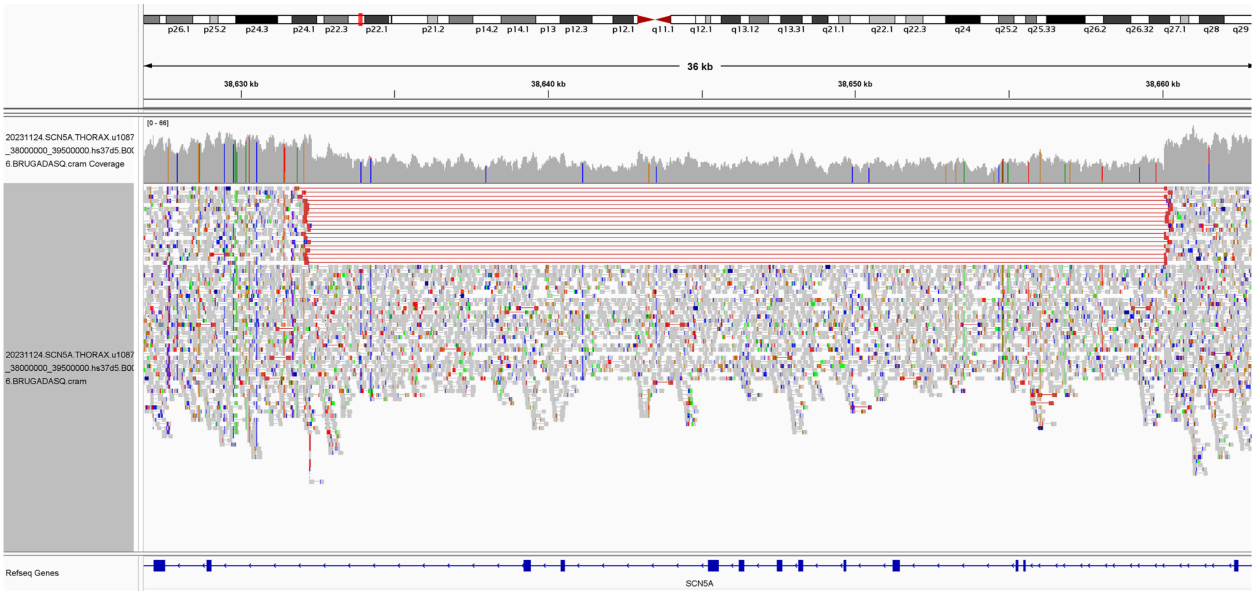


**Figure S8 | Read evidence for SV1.** A view of the Illumina paired-end WGS reads for one French BrS case in the *SCN5A* gene showing a heterozygous deletion spanning multiple exons of *SCN5A* (Integrative Genomics Viewer).

### Figure S9: Read evidence for SV2

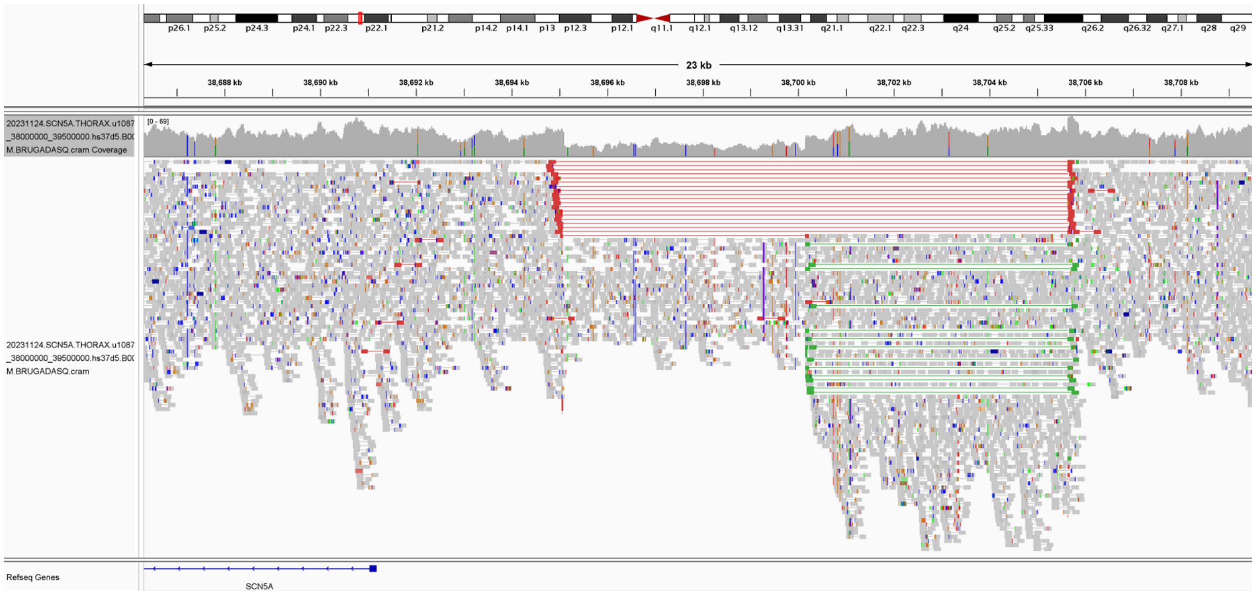


**Figure S9 | Read evidence for SV2.** A view of the Illumina paired-end WGS reads for one French BrS case upstream of the *SCN5A* gene showing a heterozygous deletion spanning an upstream CRE of *SCN5A* (Integrative Genomics Viewer).

### Figure S10: Read evidence for the mobile element insertion

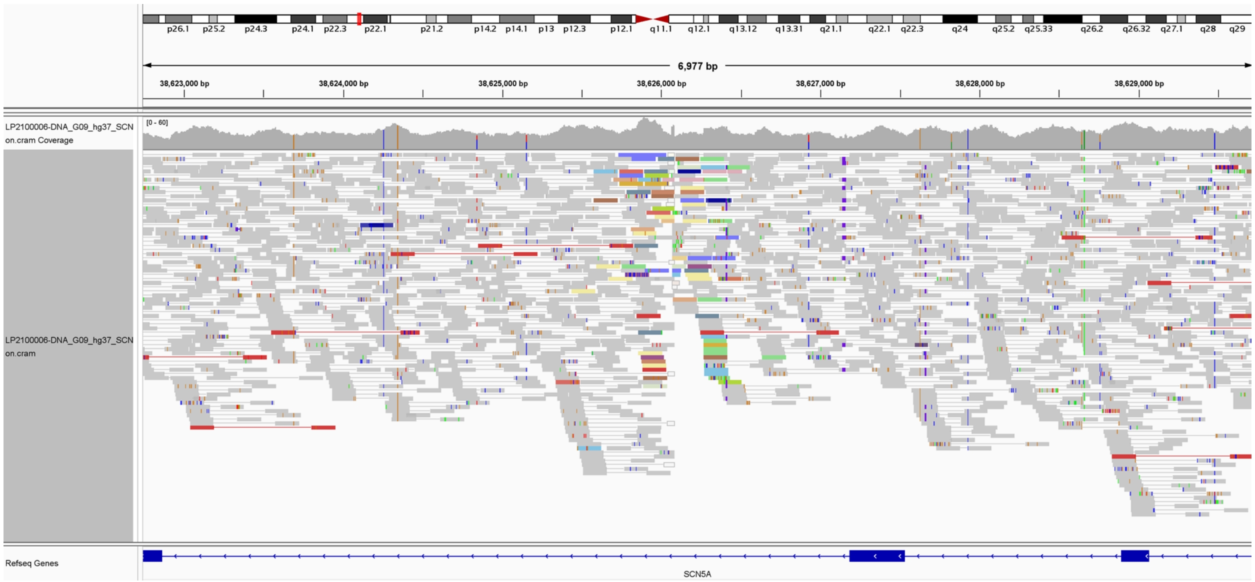


**Figure S10 | Read evidence for the mobile element insertion.** A view of the Illumina paired-end WGS reads for one Dutch BrS case in the *SCN5A* gene showing the typical signature of a retrotransposon insertion (the insertion is common, one carrier is shown here; viewed using the Integrative Genomics Viewer).

### Figure S11: Low-frequency single-variant conditional analysis

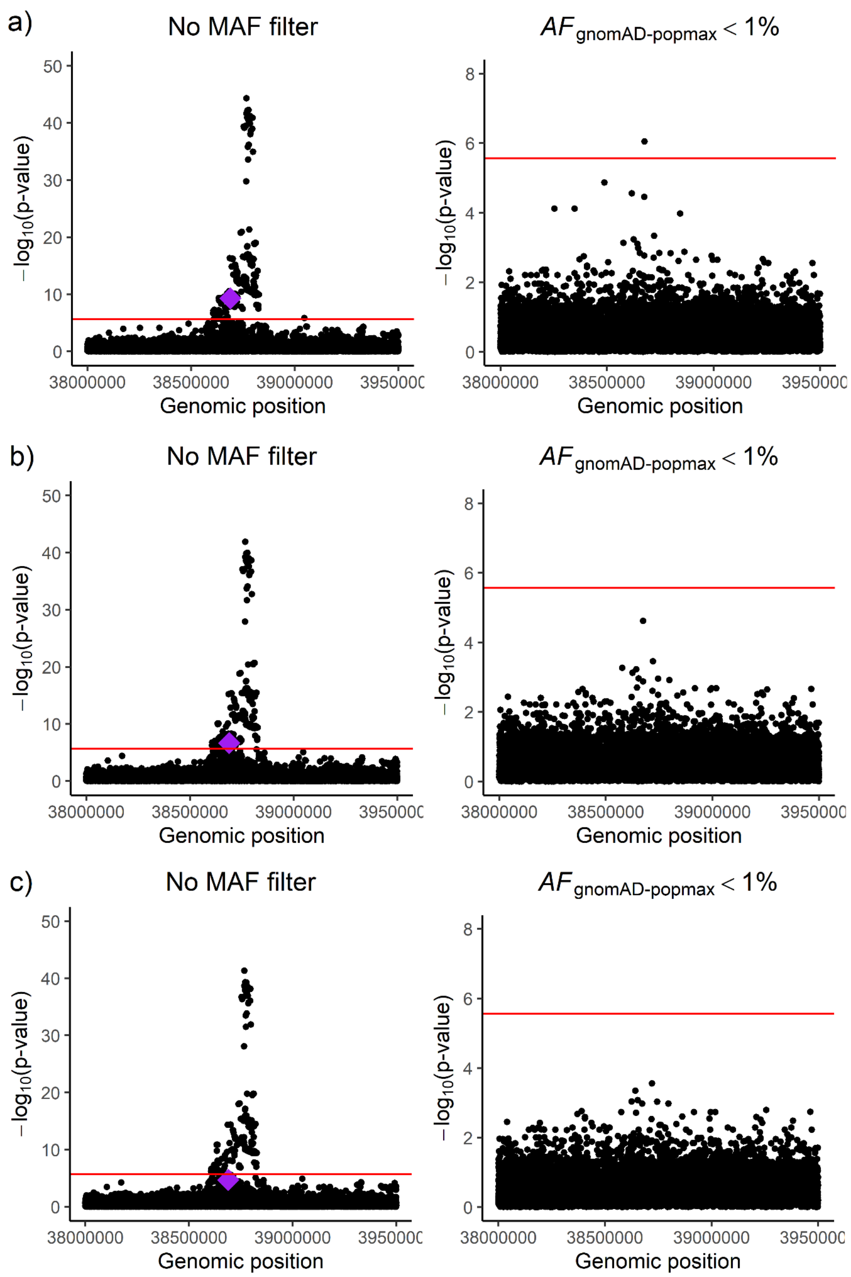


**Figure S11 | Low-frequency single-variant conditional analysis.** A) Single-variant analysis across chr3:38.0-39.5 Mb with no MAF filter (left) and with an $AF_{gnomAD\text{-}popmax}$ < 1% filter (right). Red line, Bonferroni-corrected significance level. Purple diamond, GWAS SNP rs41310232. B) Same analysis except conditioned on the lead low-frequency variant rs186741256 which is in intron 1 (3:38675954:C:A) and reduces the significance of the GWAS SNP rs41310232.

**Figure S11 |** *(Continued.)* C) Conditioned on the prior variant rs186741256 (3:38675954:C:A) and the next lead low-frequency variant which is also in intron 1 rs45527135 (3:38675003:T:G), further reducing the significance of the GWAS SNP rs41310232.
